# Non-invasive glucose monitoring vs iCGM: a systematic review and meta-analysis of accuracy and methodological challenges

**DOI:** 10.64898/2026.04.24.26351680

**Authors:** Hengrui Zhang, Ellen Dromard, Kevin C.H. Tsang, Amparo Güemes, Zhijun Guo, Stephanie E. Baldeweg, Kezhi Li

## Abstract

Non-invasive glucose monitoring (NIGM) has been pursued for decades, yet no device has achieved regulatory approval despite numerous studies reporting high accuracy. This systematic review and meta-analysis of 32 studies (38 cohorts: 20 NIGM, 18 iCGM; N = 1,693) investigated methodological factors underlying this accuracy–regulatory gap. The pooled Mean Absolute Relative Difference (MARD) for NIGM (10.21%; 95% CI: 8.73–11.69%) showed no significant difference from iCGM (11.82%; 95% CI: 10.36–13.29%; p = 0.13), with extreme heterogeneity (I² = 95.2%). Meta-regression revealed that study duration was the strongest predictor of NIGM accuracy (β = 3.94, p < 0.001), with MARD degrading from 8.7% in short-term to 15.2% in long-term studies, while iCGM accuracy remained stable. Only 15% of NIGM cohorts validated in the hypoglycemia range, compared to 89% of iCGM studies (p < 0.001). These findings suggest that reported NIGM accuracy is substantially influenced by methodological asymmetries.

## Introduction

The management of diabetes mellitus—a chronic metabolic disorder characterised by impaired glucose homeostasis, encompassing both Type 1 (autoimmune insulin deficiency) and Type 2 (insulin resistance with relative insulin deficiency) forms—represents a global health challenge affecting over 537 million adults worldwide, with projections reaching 783 million by 2045 [1]. This challenge has been transformed by the technological evolution from intermittent self-monitoring of blood glucose (SMBG) to continuous glucose monitoring (CGM) [2, 3]. In contrast to the discrete measurements provided by traditional fingerstick tests, most CGM systems provide a continuous, real-time assessment of interstitial glucose fluctuations (typically at 1 to 5-minute intervals), enabling proactive management and timely alerts for impending hypoglycemia or hyperglycemia [4, 5]. This continuous data stream has been demonstrated to improve glycemic control, reduce the risk of severe hypoglycemia, and enhance quality of life for individuals with diabetes [6, 7].

The clinical significance of accurate glucose monitoring, particularly at the extremes of the glycemic range, is substantial. Hypoglycemia can precipitate immediate cognitive impairment, seizures, loss of consciousness, and death, whereas chronic hyperglycemia constitutes the primary driver of long-term complications of diabetes including retinopathy, nephropathy, and cardiovascular disease [8]. International standards, such as ISO 15197:2013, have established minimum accuracy criteria for blood glucose monitoring systems [9]. The standard mandates that ≥95% of measured glucose values must fall within ±15 mg/dL of the reference value at concentrations <100 mg/dL and within ±15% at concentrations ≥100 mg/dL. However, the intermittent nature of SMBG leaves significant glycemic excursions undetected, highlighting the clinical need for continuous monitoring solutions [10].

The widely adopted solution for continuous glucose monitoring, particularly for individuals with type 1 diabetes, is dominated by minimally invasive CGM systems, hereafter collectively referred to as iCGM for brevity, although we note that this term has a specific regulatory meaning under FDA’s Special Controls [17]. In this review, we use iCGM broadly to encompass FDA-cleared integrated CGM devices as well as other commercially available or investigational subcutaneous and implantable sensors [11]. These devices have achieved accuracy levels that rival traditional SMBG. Leading commercial devices, including the Dexcom G6/G7, Abbott FreeStyle Libre 2/3, and the implantable Senseonics Eversense E3, consistently report a Mean Absolute Relative Difference (MARD) below the 10% threshold—an established industry benchmark for non-adjunctive use, permitting insulin dosing directly from CGM readings without confirmatory fingerstick measurements [12, 13]. Key clinical studies report an overall MARD of 8.2% for the Dexcom G7 [14], 7.8% for the Abbott FreeStyle Libre 3 [15], and 9.6% for the 90-day implantable Eversense [16]. This level of performance has led regulatory bodies such as the U.S. Food and Drug Administration (FDA) to establish a specific category for iCGM, creating a benchmark against which emerging technologies are implicitly evaluated [17]. The FDA’s iCGM special controls explicitly require that devices meet specific accuracy thresholds across the entire glycemic range, including the 15/15 criterion (≥85% of readings within 15 mg/dL or 15% of reference) and the 20/20 criterion (≥70% within 20 mg/dL or 20%) in the hypoglycemic range (<70 mg/dL) [17].

Despite these achievements, the invasive nature of current CGM technologies remains a significant barrier to wider adoption and long-term adherence. The necessity of subcutaneous sensor insertion or surgical implantation introduces challenges related to cost, discomfort, skin irritation, scarring, and infection risk [18, 19]. These limitations have sustained a decades-long pursuit of a reliable non-invasive glucose monitoring (NIGM) device capable of measuring glucose accurately and continuously without penetrating the skin [20, 21].

Non-invasive glucose monitoring encompasses a diverse array of technologies, each employing distinct physical principles to estimate blood glucose from surrounding tissues. These can be broadly categorised into four main approaches: optical, electrochemical, microwave, and electrical methods, as illustrated in Figure 1.

**Figure 1.**
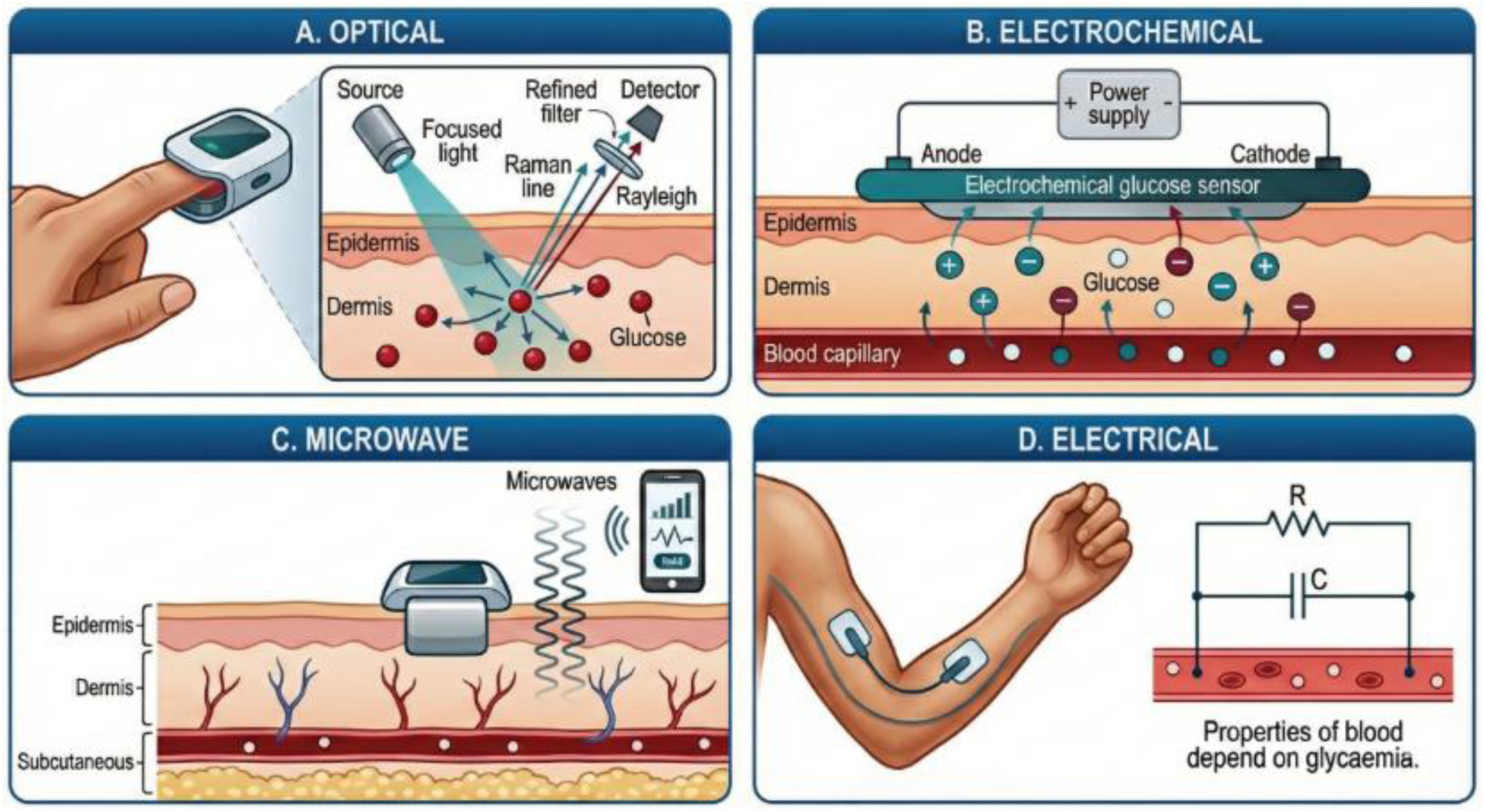
Schematic illustration of diverse sensing modalities for non-invasive glucose monitoring (NIGM). This figure outlines the fundamental operating principles of four primary categories of non-invasive sensing technologies: (A) Optical methods (e.g., Raman spectroscopy) [22, 23]; (B) Electrochemical methods (e.g., reverse iontophoresis) [24, 25]; (C) Microwave methods [26, 27]; and (D) Electrical methods (e.g., bioimpedance spectroscopy) [28, 29].

Numerous investigations have explored these technologies, including optical methods (e.g., near-infrared, Raman, and photoacoustic spectroscopy), electromagnetic approaches (e.g., microwave and radiofrequency sensing), electrochemical analysis of alternative biofluids (e.g., sweat, tears, saliva), and multimodal combinations such as ultrasonic, electromagnetic, and thermal sensing [22, 23, 30, 31, 45]. Many of these studies report MARD values below the 10% threshold, suggesting performance comparable to, or exceeding, that of established iCGM systems [37, 38, 46–49, 52, 53, 58, 59, 61, 66].

This apparent accuracy parity has created an accuracy–regulatory gap: despite numerous publications claiming high accuracy, no NIGM device has received regulatory clearance for diabetes management. This disconnect between academic claims and clinical reality was addressed in a 2024 safety communication from the FDA. The agency explicitly warned consumers: “Do not use smartwatches or smart rings that claim to measure blood glucose levels… The FDA has not authorised, cleared, or approved any smartwatch or smart ring that is intended to measure or estimate blood glucose values on its own” [32]. The FDA emphasised the life-threatening risks, noting that inaccurate measurements could lead to errors in medication dosing, potentially resulting in “mental confusion, coma, or death within hours” [32]. This warning highlights the necessity of comparing emerging NIGM devices against the established iCGM standard; if a device is intended to replace iCGM for continuous, real-time decision-making, it must be held to equivalent performance and safety standards.

This gap indicates that the accuracy reported in much of the NIGM literature might be an artifact of systemic methodological shortcomings rather than true technological maturity. Several previous reviews have surveyed the NIGM field [22, 23, 31, 33], with some focusing on specific technologies such as near-infrared spectroscopy [30] and others on particular clinical challenges such as hypoglycemia detection [35]. While Lindner et al. [35] concluded that non-invasive devices were not sufficiently accurate for detecting hypoglycemia, and other narrative reviews have highlighted general challenges [36], to our knowledge, no systematic review has quantitatively examined the specific methodological factors that might contribute to less reliable accuracy estimates across the entire field.

This systematic review and meta-analysis aims to address this critical gap. We hypothesise that the apparent accuracy of NIGM devices might be systematically overestimated due to three methodological biases: (1) the use of lower-precision reference standards that introduce comparator noise and render true device error unverifiable; (2) the reliance on short-duration validation protocols that fail to capture long-term sensor drift; and (3) the systematic avoidance of testing in the hypoglycemic range. By quantifying the impact of these methodological asymmetries, we seek to resolve the accuracy–regulatory gap, provide an evidence-based explanation for the field’s regulatory challenges, and establish a rigorous framework for the clinical validation of future NIGM technologies.

## Results

### Study selection and characteristics

The systematic search and selection process is detailed in the PRISMA 2020 flow diagram (Figure 2). Our search identified 1,382 records from three databases. After removing duplicates and screening titles and abstracts, 47 full-text articles were assessed for eligibility. Following application of inclusion and exclusion criteria, 32 studies comprising 38 distinct cohorts were included in the final analysis. Studies were excluded primarily for the following reasons: Lack of application to glucose monitoring in diabetes (n = 2), change and/or difference in MARD (n = 4) and computer simulated or non-primary data (n = 8), and insufficient data (n = 1). The full list of included studies is provided in the References section [37–44, 46–69].

**Figure 2.**
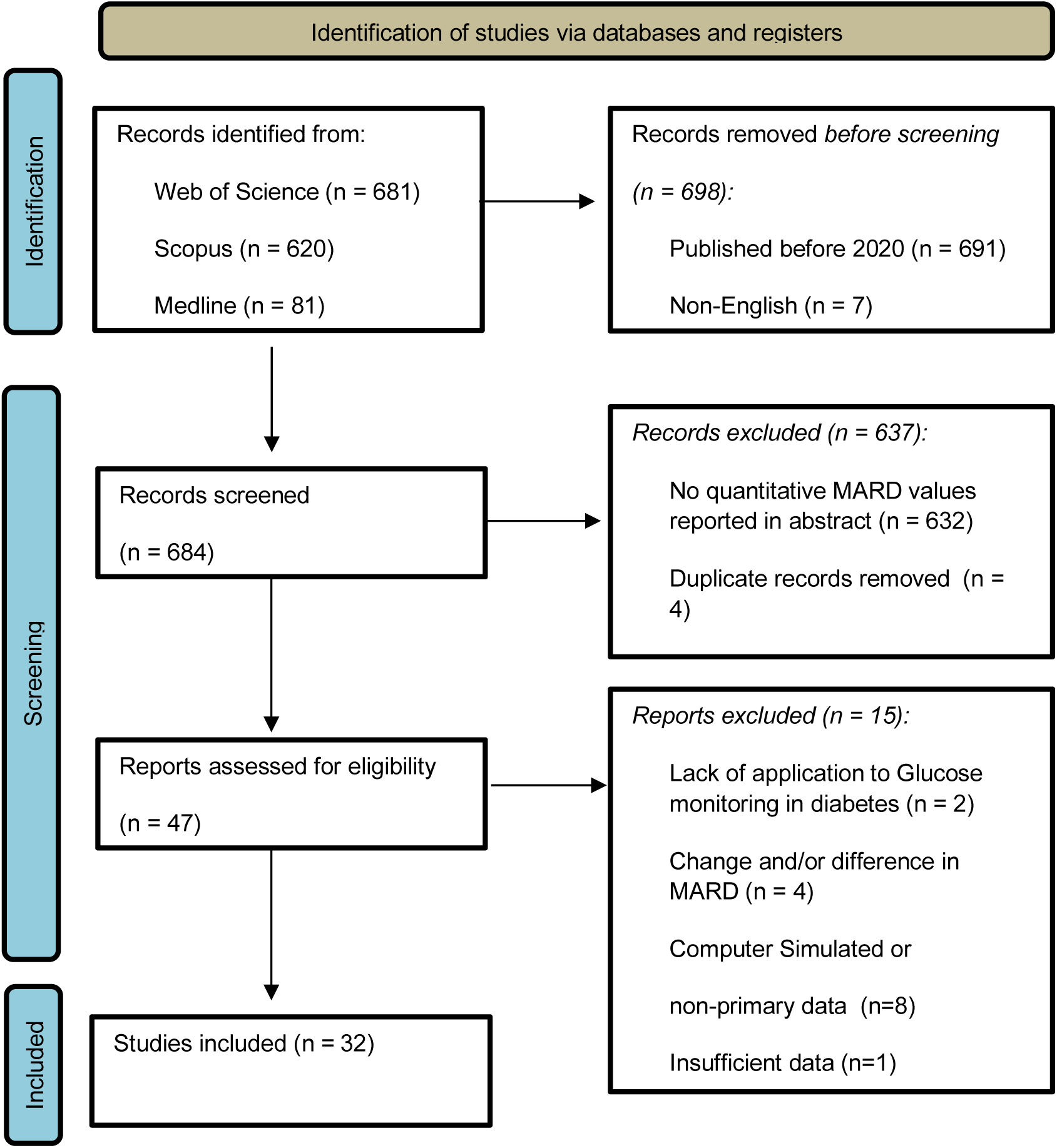
PRISMA 2020 Flow Diagram. The systematic search identified 1,382 records, with 32 studies (38 cohorts) meeting inclusion criteria after screening and eligibility assessment.

The 38 cohorts included 20 NIGM cohorts (N = 981) and 18 iCGM cohorts (N = 712), with a total of 1,693 participants [37–44,46–69]. Publication years ranged from 2020 to 2025. Study durations ranged from single measurements to 180 days, with a median of 4 hours for NIGM studies and 14 days for iCGM studies. Sample sizes ranged from 1 to 187 participants per cohort (note: one cohort, Suminaga et al. 2020, comprised a single participant; a sensitivity analysis excluding this study did not materially alter the pooled estimates).

Table 1 presents a summary of the key audit characteristics across device categories, highlighting the fundamental methodological asymmetries between NIGM and iCGM validation studies (Figure 3).

**Figure 3.**
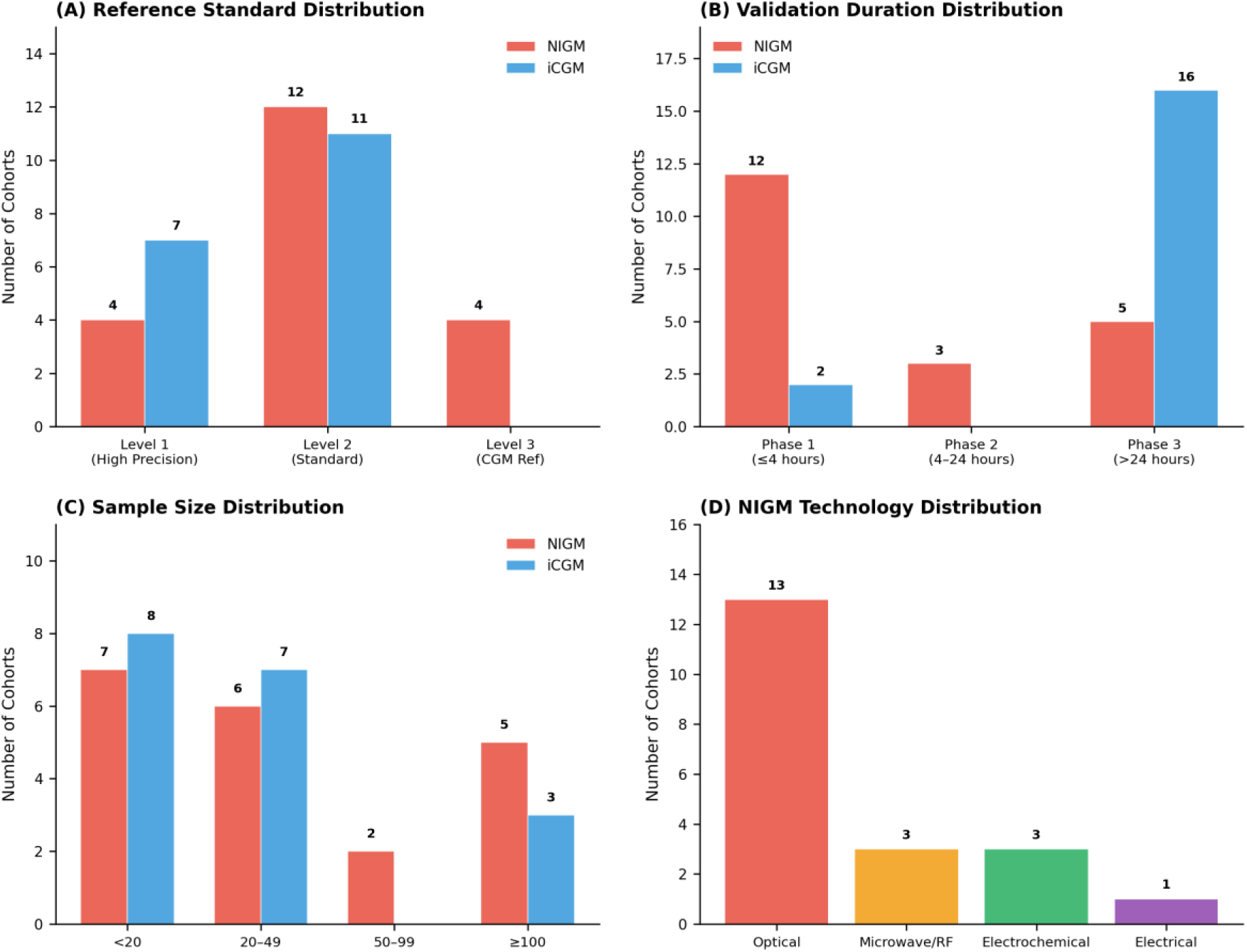
Methodological Characteristics of Included Cohorts. Bar charts showing the distribution of (A) reference standards, (B) validation durations, (C) sample sizes, and (D) NIGM technology types for NIGM and iCGM cohorts.

**Table 1.**
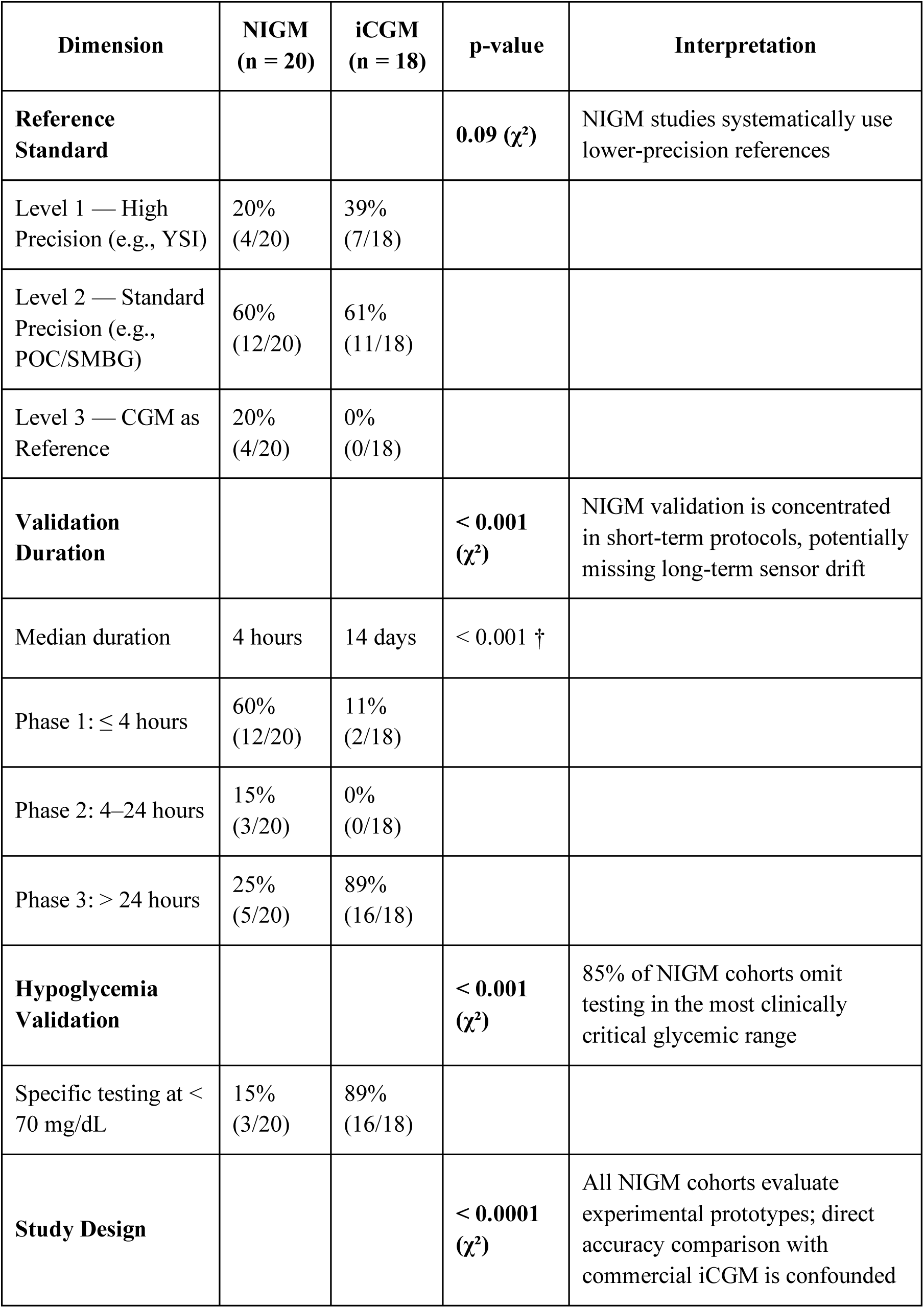
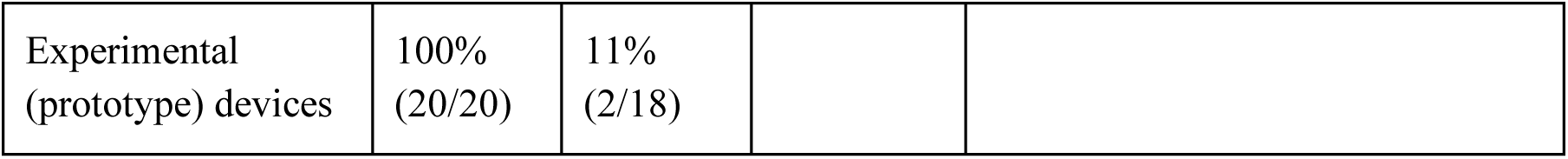
Summary Table: Methodological Characteristics by Device Category. The complete characteristics of all 38 included cohorts are provided in Supplementary Table S5.

For iCGM devices, a numerical difference in the choice of reference standards was observed; meta-regression indicated a trend toward higher MARD with lower-quality references (β = 4.55 percentage points per level decrease), although this did not reach statistical significance (p = 0.09; Supplementary Figure S2). No such trend was observed for NIGM devices (p = 0.49). Only 20% (4/20) of NIGM cohorts used Level 1 high-precision references (e.g., YSI analyser), compared to 39% (7/18) of iCGM cohorts. The majority of NIGM studies relied on Level 2 (60%) or Level 3 (20%) references, which tend to produce less reliable accuracy estimates by introducing measurement noise in the comparator.

All 20 NIGM cohorts evaluated experimental prototype devices, whereas only 11% (2/18) of iCGM cohorts used experimental systems (p < 0.0001). The remaining 89% of iCGM studies validated established, regulatory-approved commercial products. This fundamental asymmetry in technological maturity represents a major confounding factor in any direct comparison of reported accuracy metrics.

### Pooled accuracy estimates

The random-effects meta-analysis yielded a pooled MARD of 10.96% (95% CI: 9.89–12.02%) across all 38 cohorts (Figure 4). Heterogeneity was extreme (Cochran’s Q, df = 37, p < 0.001; I² = 95.2%, τ² = 9.54), indicating that the true effect size varies substantially across studies. When stratified by device type, the pooled MARD for NIGM devices (10.21%; 95% CI: 8.73–11.69%) was not significantly different from that of iCGM devices (11.82%; 95% CI: 10.36–13.29%; p = 0.13). That NIGM devices—none of which have achieved regulatory approval—appear numerically more accurate than established iCGM systems is suggestive of the methodological biases explored in subsequent subgroup analyses.

**Figure 4.**
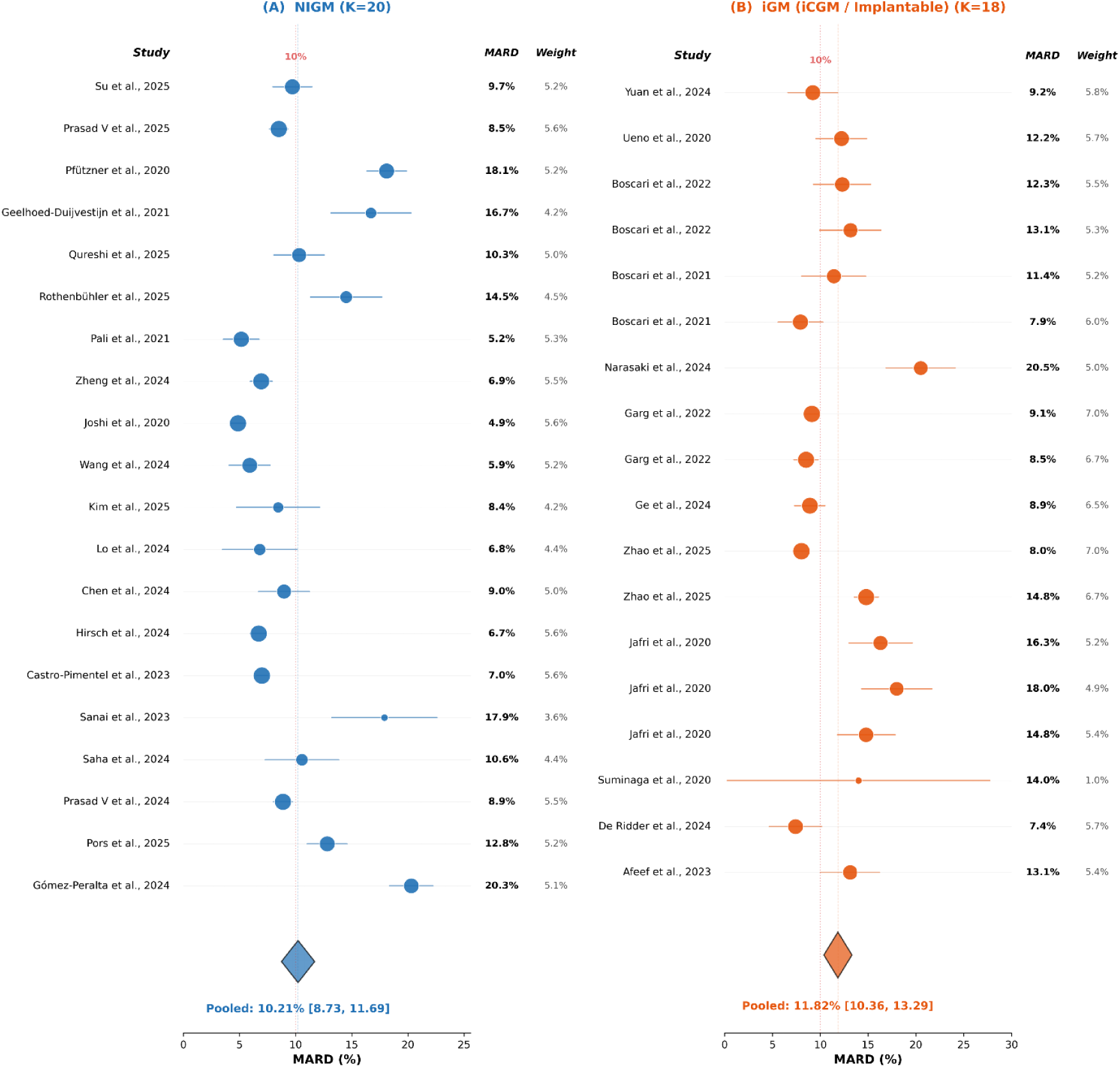
Forest Plot of MARD Estimates. Random-effects meta-analysis of 38 cohorts showing an overall pooled MARD of 10.96% (95% CI: 9.89–12.02%) with extreme heterogeneity (I² = 95.2%). Subgroup analysis yielded pooled MARD of 10.21% (95% CI: 8.73–11.69%) for NIGM (K=20) and 11.82% (95% CI: 10.36–13.29%) for iCGM

### Duration–accuracy relationship

Meta-regression analysis revealed a significant interaction between device type and study duration (Figure 5). The temporal scope of validation differed dramatically between the two groups: the median study duration for NIGM was 4 hours versus 14 days for iCGM (p < 0.001), with 60% (12/20) of NIGM cohorts conducting validation over ≤4 hours compared to only 11% (2/18) of iCGM cohorts, and 89% (16/18) of iCGM studies extending beyond 24 hours versus just 25% (5/20) of NIGM studies. This concentration of NIGM validation in short-duration protocols might systematically fail to capture sensor drift, calibration decay, and performance degradation over time. Separate meta-regressions showed that for NIGM devices, longer study durations were associated with significantly higher MARD values (β = 3.94, p < 0.001, R² = 0.32), with accuracy degrading from approximately 8.7% in Phase 1 (≤4 hours) to 15.2% in Phase 3 (>24 hours). In contrast, iCGM devices maintained stable performance regardless of study duration (β = 0.19, p = 0.78), with MARD ranging from 11.4% in Phase 1 to 12.3% in Phase 3. The interaction term (Duration × Device Type) in the full model confirmed this differential effect, with NIGM devices showing significantly greater accuracy degradation over time compared to iCGM devices.

**Figure 5.**
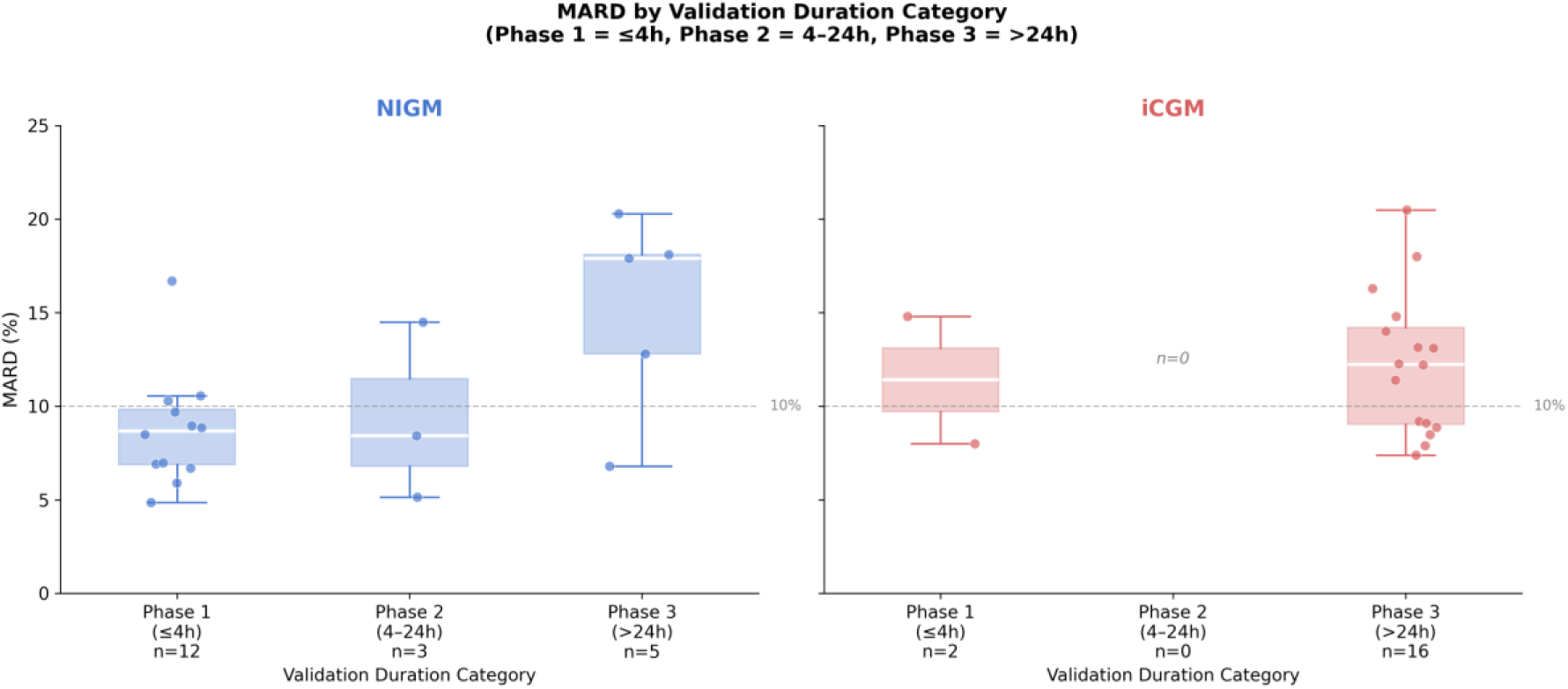
Duration-MARD Analysis. (A) Scatter plot showing a significant positive association between validation duration and MARD for NIGM (β = 3.94, p < 0.001) but not for iCGM (β = 0.19, p = 0.78). Point size is proportional to sample size. (B) Bar chart showing subgroup MARD by duration phase, highlighting the degradation of NIGM accuracy in long-term studies.

### Hypoglycemia validation

Analysis of hypoglycemia validation revealed a critical gap in the NIGM literature (Figure 6). Of the 20 NIGM cohorts, only 3 (15%) included specific testing in the hypoglycemic range (<70 mg/dL), compared to 16 of 18 (89%) iCGM cohorts (p < 0.001). This represents a systematic gap in the evaluation of device performance in the most clinically critical and potentially life-threatening scenarios.

**Figure 6.**
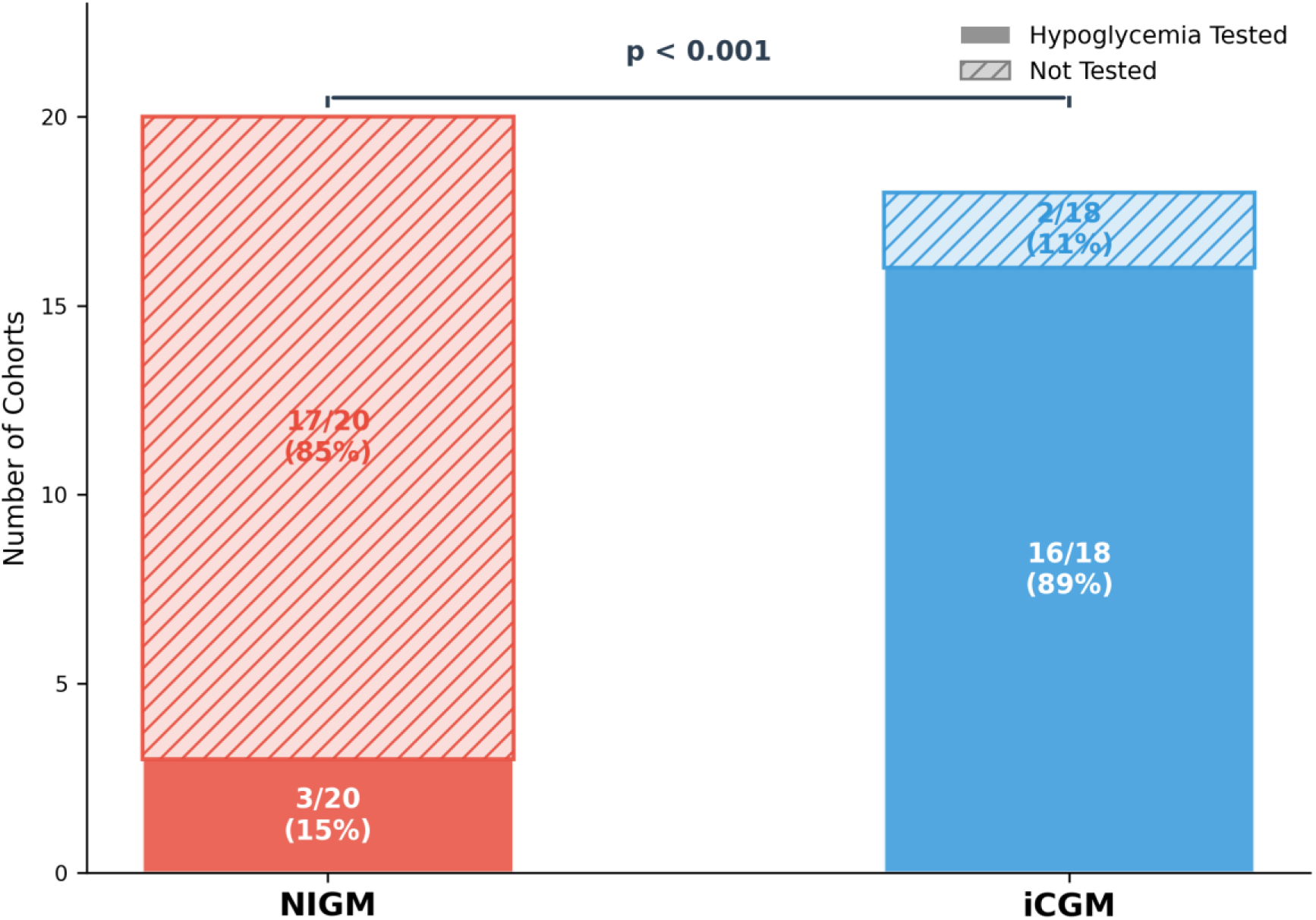
Hypoglycemia Validation Gap. Stacked bar chart showing the proportion of NIGM and iCGM cohorts that included specific testing in the hypoglycemic range (<70 mg/dL). A significantly smaller proportion of NIGM studies performed this critical safety testing (15% vs 89%, p < 0.001).

### Rishk of bias assessment

Quality assessment using the QUADAS-2 tool revealed substantial risk of bias across the included studies, displayed in NIGM and iCGM categories (Figure 7). NIGM studies exhibited higher risk of bias, with 55% (11/20) rated as high risk in at least one domain, compared to 6% (1/18) for iCGM. The inter-rater reliability for the QUADAS-2 assessment was excellent (ICC = 0.85, 95% CI: 0.80–0.89). The complete QUADAS-2 assessment with rationale is provided in Supplementary Table S2.

**Figure 7.**
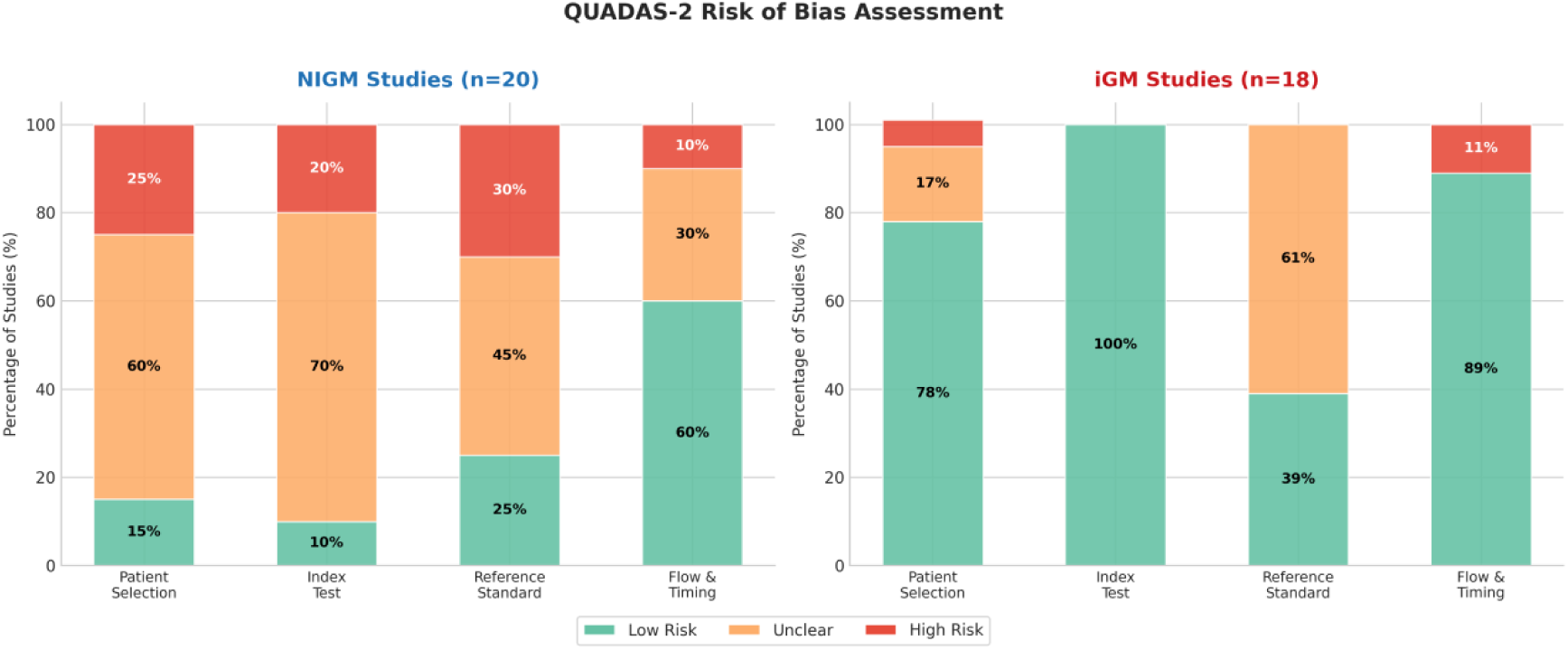
QUADAS-2 Risk of Bias Summary for NIGM and iCGM Studies. Distribution of risk of bias across four QUADAS-2 domains (Patient Selection, Index Test, Reference Standard, Flow & Timing) for the 20 NIGM cohorts and 18 iCGM cohorts. NIGM studies exhibited substantially higher proportions of high-risk ratings across all domains compared to iCGM studies.

### Sensitivity analysis and publication bias

Leave-one-out sensitivity analysis demonstrated that the pooled MARD estimate was consistent, with values ranging from 10.63% to 11.14% (Supplementary Figure S1 and Supplementary Table S4). No single study had a disproportionate influence on the overall result.

Egger’s test indicated significant asymmetry (p = 0.0002), and Begg’s test was also significant (τ = 0.41, p = 0.0003), suggesting the presence of publication bias (Supplementary Figure S3). This asymmetry is consistent with the selective non-publication of studies reporting higher (less favourable) MARD values, implying that the pooled estimates presented here may be optimistically biased.

### Certainty of evidence

Using the “Grading of Recommendations Assessment, Development and Evaluation” (GRADE) framework, the overall certainty of evidence for NIGM accuracy was rated as Very Low in an exploratory adapted GRADE assessment due to: (1) high risk of bias in the majority of studies; (2) very serious inconsistency (I² = 95.2%); (3) serious indirectness (most studies used healthy volunteers rather than the target population); (4) serious imprecision (wide confidence intervals reflecting small cumulative sample sizes and extreme heterogeneity); and (5) high suspicion of publication bias. The complete GRADE assessment is provided in Supplementary Table S3.

### Technology-stratified analysis

To examine whether MARD performance varies systematically across NIGM technology categories, we stratified the 20 NIGM cohorts by their primary sensing modality (Figure 8). Optical methods (including NIR spectroscopy and multi-modal approaches) showed the highest variability. However, these results should be interpreted with caution given the small number of cohorts within each technology category, which limits the statistical power of between-group comparisons.

**Figure 8.**
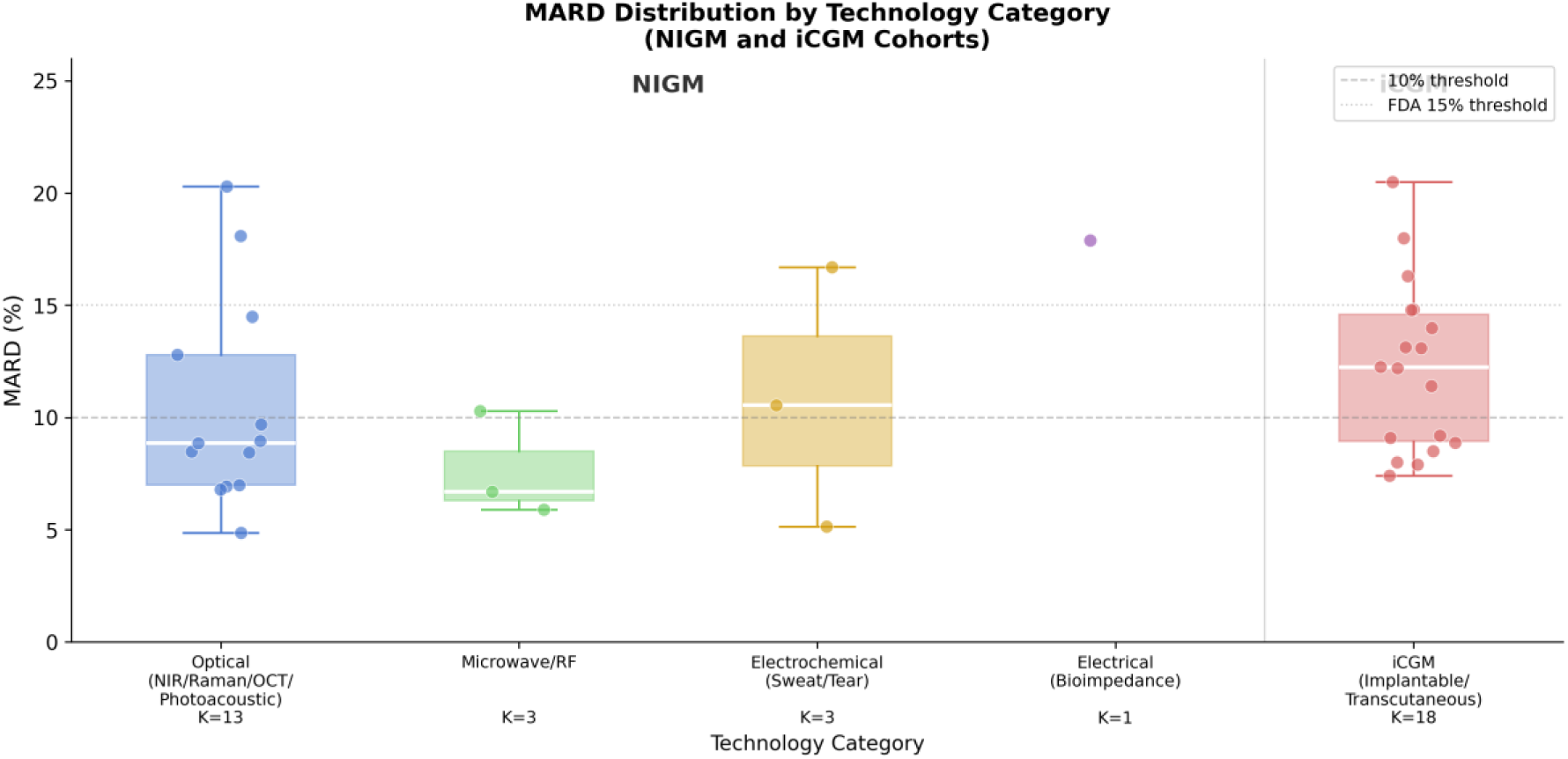
Technology-Stratified MARD Distribution for NIGM Devices. Box plots showing the distribution of reported MARD values across four NIGM technology categories. Optical approaches showed higher variability. Each point represents one cohort.

## Discussion

To our knowledge, this is the first systematic review and meta-analysis to quantitatively examine the methodological factors that might contribute to overestimated accuracy claims in the non-invasive glucose monitoring literature. Our findings provide a data-driven explanation for the “accuracy–regulatory gap”—the persistent gap between the impressive accuracy reported in academic studies and the failure of any NIGM device to achieve regulatory approval for diabetes management [35, 88]. The pooled MARD for NIGM devices (10.21%; 95% CI: 8.73–11.69%) was not significantly different from that of iCGM devices (11.82%; 95% CI: 10.36–13.29%; p = 0.13), a paradoxical finding given that no NIGM device has achieved regulatory approval. This apparent parity, combined with extreme heterogeneity (I² = 95.2%) and the finding that 55% of NIGM cohorts were rated as high risk of bias on QUADAS-2 assessment (compared to 6% for iCGM), suggests that the reported accuracy of NIGM devices is inflated by systematic methodological shortcomings rather than reflecting genuine technological maturity.

The primary finding of this analysis is the differential sensitivity of NIGM and iCGM devices to methodological factors. For NIGM devices, study duration emerged as the dominant predictor of reported accuracy, a finding discussed in detail below. In contrast, the reference standard category was not a significant predictor of NIGM accuracy (p = 0.49), suggesting that the dominant sources of measurement variability in NIGM studies lie elsewhere—most likely in the biophysical challenges of non-invasive sensing discussed below. For iCGM devices, the reference standard category showed a numerical association with MARD (β = 4.55, p = 0.09), with Level 1 references associated with a lower mean MARD (9.4%) compared to Level 2 references (14.0%); however, this association did not reach conventional statistical significance and should be interpreted cautiously given the limited sample size.

Our analysis provides quantitative evidence for a duration-dependent bias, where the accuracy of NIGM devices significantly degrades in studies lasting longer than 24 hours. The significant association between study duration and NIGM accuracy (β = 3.94, p < 0.001, R² = 0.32) indicates that study duration explains approximately one-third of the between-study variance in NIGM MARD, with the remaining variance likely attributable to differences in technology type, calibration approach, population characteristics, and other unmeasured factors. This finding suggests that while iCGM devices maintain stable performance over weeks or months, NIGM technologies struggle to maintain signal integrity over extended periods. This finding is critical because the primary value proposition of CGM is its continuous, long-term nature. A device that is only accurate for a few hours does not constitute a true CGM replacement [22, 89, 91].

These findings likely reflect the well-documented biophysical challenges of non-invasive sensing. Unlike subcutaneous iCGM sensors, which operate in a relatively stable tissue environment, NIGM sensors are exposed to the dynamic skin surface. The stratum corneum undergoes continuous hydration and dehydration cycles driven by ambient humidity, sweating, and transepidermal water loss [70]. Water absorption in the near-infrared spectrum (at 1450 nm and 1940 nm) overlaps with the weaker glucose absorption bands [72] [73], and hydration changes similarly affect Raman-based measurements [71]. A change in skin hydration of just 10% can alter the measured optical signal by an amount equivalent to a 50–100 mg/dL change in glucose concentration [70]. Skin temperature also fluctuates with ambient conditions and circadian rhythms, altering optical properties through changes in blood perfusion [92] and the dielectric properties measured by microwave sensors [74].

A further challenge is the inherently weak glucose signal: the specific absorption coefficient of glucose in the NIR is approximately 10,000 times weaker than that of water [75], making it difficult to isolate from confounding signals. Over time, the accumulation of sebum, dead skin cells, and environmental contaminants on the sensor-skin interface further attenuates this signal. For electrochemical sensors relying on sweat, the composition varies significantly with activity level, introducing confounding signals [70]. The mechanical interface between sensor and skin also degrades over time, as tissue pressure causes local changes in blood perfusion and micro-movements alter the optical path length or electrical contact impedance [91].

These biophysical factors, which are negligible in short, controlled laboratory sessions, become dominant sources of error in real-world, long-term use. The significant Duration × Device Type differential (p < 0.001) in our meta-regression provides strong statistical evidence that these mechanisms are not merely theoretical concerns but are likely contributing to the observed performance degradation of NIGM devices in practice.

From a clinical safety perspective, our most concerning finding is the systematic avoidance of hypoglycemia testing in the NIGM literature. Our analysis showed that 85% (17/20) of NIGM cohorts failed to perform specific validation in the hypoglycemic range (<70 mg/dL), a stark contrast to the 89% (16/18) of iCGM cohorts that did. This is not merely a methodological limitation; it represents a critical gap in the evaluation of device performance in the most life-threatening scenarios. Unrecognised hypoglycemia can lead to catastrophic consequences, including seizures, coma, and death [76, 77].

The physiological response to hypoglycemia itself creates multiple confounding factors for non-invasive sensors. The counter-regulatory hormonal response triggers sweating, alterations in peripheral circulation (leading to pale, cool skin), and tremor—all of which can dramatically alter the signals measured by optical, electrical, and thermal sensors [78]. By failing to induce and validate during controlled hypoglycemic clamps, the vast majority of NIGM studies are effectively ignoring the most challenging and clinically important aspect of glucose monitoring. The FDA’s iCGM guidance explicitly requires consistent performance in the hypoglycemic range, and the inability of the NIGM field to demonstrate this basic safety prerequisite represents a major barrier to regulatory approval.

An important caveat to the NIGM–iCGM comparison concerns the developmental maturity of the devices studied. Our data show that 100% of NIGM cohorts evaluated experimental prototypes, whereas 89% of iCGM cohorts used established commercial products (p < 0.0001). This represents an unequal comparison. When we properly stratify by developmental maturity, the interpretation changes significantly. This asymmetry in technological maturity is a critical confounding factor. While a direct comparison between the 20 experimental NIGM cohorts and the 2 experimental iCGM cohorts showed no statistical difference in MARD (p = 0.87), this comparison is severely limited by the small sample size and should be interpreted with extreme caution. The more salient point is that the entire body of NIGM literature is based on prototype performance. Experimental prototypes are often validated under idealised conditions, on small homogenous populations, and may benefit from post-hoc algorithm tuning not feasible in a locked-down commercial product [31, 93].

In other words, the current literature is largely comparing early-stage prototypes against market-proven medical devices, and the apparent accuracy parity should be interpreted in this light. The challenge for NIGM is not merely to match the MARD of commercial iCGM in a controlled lab setting, but to advance from a promising prototype to a regulatory-approved system that maintains its performance in the complexities of real-world clinical use.

Resolving the accuracy–regulatory gap requires moving beyond the simplistic goal of achieving a low MARD and instead adopting a differentiated, application-driven approach to technology development and validation. The evidence from our analysis suggests that the assumption of a single, universal “NIGM” device might be overly simplistic. Instead, the field should pursue three distinct, evidence-based pathways, each with its own set of performance requirements, clinical use cases, and standardised, generalisable validation methods and metrics, a vision supported by the growing integration of AI and wearable technology in diabetes management [88].

The first pathway is for technologies aiming to replace current iCGM systems for full-time, non-adjunctive use. The performance bar is exceptionally high: MARD < 10% (ideally < 8%) validated against a Level 1 reference, proven stability over at least 7–14 days, and consistent accuracy in the hypoglycemic range. As shown in the technology-stratified analysis (Figure 8), our analysis suggests that technologies with deeper tissue penetration and less susceptibility to skin surface variability, such as Microwave/RF sensing [59, 74] and potentially implantable optical sensors, are the most promising candidates for this pathway.

The second pathway is for technologies designed for intermittent, on-demand glucose checks, aiming to replace the discomfort of traditional fingersticks. The requirement for long-term stability is relaxed, but the demand for high accuracy in a single measurement is essential. Technologies that can provide a rapid, accurate “metabolic snapshot,” such as photoacoustic spectroscopy [34, 38, 79] and advanced Raman spectroscopy systems [43, 80], are well-suited for this niche.

The third pathway moves away from a sole focus on medical-grade glucose accuracy and instead embraces a broader vision of metabolic health monitoring. Rather than pursuing the stringent accuracy required for clinical decision such as insulin dosing, these devices aim to provide metabolic trend awareness and lifestyle guidance for the growing population of health-conscious consumers and individuals at risk of metabolic syndrome. This approach integrates moderate-accuracy glucose sensing with other physiological parameters (e.g., heart rate, activity, skin temperature, lactate) [89, 90] to deliver integrated wellness insights. The commercial viability of this pathway is increasingly evident, with emerging technologies leveraging exhaled breath analysis, photoplethysmography (PPG), and multi-analyte microneedle sensors to provide metabolic trend awareness rather than medical-grade accuracy [81, 100, 101]. Critically, the regulatory landscape is evolving to accommodate this paradigm. The U.S. FDA’s updated General Wellness guidance (2026) permits devices derived from blood glucose measurements to operate outside the medical device regulatory pathway when marketed for general wellness purposes, provided they do not make diagnostic claims [102]. This regulatory shift may accelerate the development and commercialisation of wellness-oriented glucose monitoring technologies, potentially expanding the addressable market well beyond the population with diabetes.

From a policy perspective, our findings suggest that regulatory bodies should consider establishing a tiered framework for the evaluation of glucose monitoring technologies, with distinct requirements tailored to the intended use of the device. For devices intended for therapeutic decision-making, the current rigorous iCGM standards should be maintained. For point-of-care devices, standards could focus on single-point accuracy against a Level 1 reference. For wellness devices, the focus could shift from MARD to metrics that capture trend accuracy and the ability to detect meaningful glycemic variability, without the stringent requirements for hypoglycemic alarms [31, 93].

The overall certainty of evidence, assessed using an adapted GRADE framework, was rated as Very Low, reflecting the cumulative impact of high risk of bias, extreme heterogeneity, serious indirectness, imprecision, and suspected publication bias. We note that the standard GRADE framework was designed for intervention effects and diagnostic test accuracy; its application to pooled MARD is exploratory, and QUADAS-2 serves as the primary quality assessment tool in this review. This rating indicates that the true accuracy of NIGM devices may differ substantially from the pooled estimates reported here, and further research is very likely to change the estimate. This study has several limitations that warrant consideration. First, the high heterogeneity (I² = 95.2%) across studies, while itself a key finding, limits the precision of the pooled estimates and means the summary MARD should be interpreted as an average of a wide range of effects rather than a single true value. As Borenstein [94] notes, I² reflects the proportion of variance due to heterogeneity rather than its absolute magnitude, and high heterogeneity is a well-documented and expected feature of meta-analyses involving continuous outcomes [95, 96]. However, the primary goal of this study was not to derive a precise point estimate, but to use meta-regression to explain this very heterogeneity. This heterogeneity is therefore an inherent feature of a field with such diverse technologies and methodologies. We used the DerSimonian-Laird estimator for the primary analysis, which is known to underestimate between-study variance when heterogeneity is high. A sensitivity analysis using the restricted maximum likelihood (REML) estimator yielded similar results (Supplementary Table S4), supporting the robustness of our findings. Second, the presence of significant publication bias, as indicated by Egger’s test (p = 0.0002), suggests that our pooled MARD estimates may be optimistic, as studies with less favourable results are likely underreported. Third, our analysis relied on study-level data, which prevented a more granular, patient-level analysis of factors influencing accuracy. Fourth, this review did not analyse the potential impact of participant skin tone (e.g., Fitzpatrick skin type) as a confounding variable, which is a recognised limitation particularly for optical sensing technologies, and an important consideration for health equity in future studies. Finally, our search was limited to studies published since 2020 to capture the contemporary field, which may have excluded some earlier, foundational work.

This systematic review and meta-analysis provides quantitative evidence suggesting that the “accuracy–regulatory gap” in non-invasive glucose monitoring is strongly associated with, and likely influenced by, systemic methodological limitations in the published literature. Of the three a priori hypotheses tested, the duration-dependent bias hypothesis received the strongest support (β = 3.94, p < 0.001), demonstrating that NIGM accuracy degrades significantly in studies lasting longer than 24 hours. The hypoglycemia testing hypothesis was also strongly supported, with 85% of NIGM cohorts failing to validate in this clinically critical range. The reference standard hypothesis, while showing a numerical trend for iCGM devices (β = 4.55, p = 0.09), did not reach statistical significance. Collectively, these findings indicate that the impressive accuracy reported by many NIGM studies appears to be substantially influenced by the reliance on short validation periods and the avoidance of hypoglycemia testing. The future of NIGM is not a single device but a portfolio of technologies tailored to specific clinical needs. By embracing a rigorous, safety-first validation framework and pursuing a differentiated, application-driven roadmap, the field can finally move beyond the pattern of overpromising and begin to achieve the goal of truly non-invasive glucose monitoring.

## Methods

### Study registration and protocol

This systematic review and meta-analysis was conducted in accordance with the PRISMA 2020 statement [82]. This review was registered in PROSPERO (CRD420261325372). The registration was submitted after the commencement of data extraction but prior to the completion of the analysis. A formal protocol was not prepared; however, the analytical framework, including the pre-specified audit criteria (Table 2), was established prior to data extraction. The PRISMA 2020 checklist is provided in Supplementary Table S6.

**Table 2.** Audit Specification Table.

| Dimension | Level/Phase | Definition | Rationale |
| --- | --- | --- | --- |
| **Reference Standard Quality** | Level 1 (High Precision) | Laboratory-grade analyzers (YSI, EKF Biosen, Beckman); CV < 2% | Gold standard for pivotal trials |
|  | Level 2 (Standard Precision) | Hospital-grade POC or high-quality SMBG; CV 2–5% | Acceptable for clinical use |
|  | Level 3 (CGM Reference) | Another CGM device as reference; CV 5–10% | Circular validation; inflates agreement |
| **Validation Duration** | Phase 1 (Short-term) | ≤4 hours; Single session | May miss drift and confounders |
|  | Phase 2 (Medium-term) | 4–24 hours; Extended session | Captures some physiological variation |
|  | Phase 3 (Long-term) | >24 hours; Multi-day wear | Captures real-world conditions |
| **Safety Test** | Hypoglycemia Testing | Specific validation at <70 mg/dL | Essential for clinical use approval |

### Search strategy

We performed a systematic search of three electronic databases—Web of Science (Core Collection), Medline (via PubMed), and Scopus—for studies published between January 1, 2020, and November 30, 2025. The search strategy combined terms related to glucose monitoring devices (e.g., “CGM,” “glucometer,” “glucose monitor”), technology types (e.g., “non-invasive,” “implantable,” “transdermal,” “biosensors”), and the condition (e.g., “diabet,” “T1D,” “T2D”). A NOT operator was applied to exclude animal and in vitro studies at the search level. The full search strategy for each database is provided in the Supplementary Materials (S1). The electronic database search was supplemented by a manual review of reference lists from included articles and relevant systematic reviews, as well as papers identified during a preliminary pilot study, to ensure broad coverage.

### Eligibility criteria

Studies were included if they met the following criteria: (1) evaluated a non-invasive or minimally invasive glucose monitoring device in humans; (2) reported the MARD as an accuracy metric; (3) were published in English; and (4) were original research articles. We excluded reviews, conference abstracts, and studies conducted solely on animals or in vitro. We also excluded studies that used computer-generated or simulated data, as well as those conducted in highly specific, non-generalisable measurement environments (e.g., during dialysis) without broader validation.

### Study selection and data extraction

Two reviewers independently screened titles and abstracts, achieving excellent inter-rater reliability (Cohen’s kappa = 0.92), and any discrepancies were resolved by consensus. The full texts of potentially eligible articles were then retrieved and assessed against the inclusion criteria. Data extraction was performed independently by two reviewers (H.Z. and E.D.) using a standardised template. Discrepancies were resolved through discussion and consensus, with a third reviewer (K.L.) consulted when necessary.

### Methodological audit framework

To systematically evaluate the methodological rigor of included studies, we developed a novel, pre-specified audit framework comprising three dimensions: Reference Standard Quality, Validation Duration, and Hypoglycemia Testing. This framework was designed *a priori* to identify the specific methodological factors, including temporal resolution of validation protocols, hypothesised to produce unreliable accuracy estimates in the NIGM literature. The complete audit specification is presented in Table 2. This framework was developed specifically for this study to systematically quantify methodological differences between the two bodies of literature. A fourth domain, time resolution (i.e., the frequency of NIGM measurements), was also considered but not formally included in the primary analysis due to inconsistent reporting across studies.

Reference standard quality was classified into three levels based on the analytical precision of the comparator device. Level 1 (High Precision) included laboratory-grade analysers such as the YSI 2300/2500 STAT Plus (Yellow Springs Instruments, OH, USA), EKF Biosen (EKF Diagnostics, Germany), and Beckman Coulter AU series (Beckman Coulter, CA, USA), which have a coefficient of variation (CV) typically < 2% [97]. Level 2 (Standard Precision) included hospital-grade point-of-care (POC) devices and high-quality self-monitoring blood glucose (SMBG) meters with a CV of 2–5%. Level 3 (CGM Reference) included studies that used another CGM device (e.g., Dexcom G6) as the reference standard, which introduces the risk of circular validation and spuriously elevated agreement due to correlated errors.

Validation duration was categorised into three phases: Phase 1 (≤4 hours), representing single-session laboratory studies; Phase 2 (4–24 hours), representing extended monitoring sessions; and Phase 3 (>24 hours), representing multi-day real-world validation. The rationale for this classification is that short-term studies might fail to capture the long-term sensor drift and physiological confounders that affect real-world performance [98].

All interaction terms (Duration × Device Type) were defined *a priori* based on biophysical drift hypotheses, specifically the expectation that non-invasive sensors would show greater accuracy degradation over time due to skin hydration cycles, thermal equilibrium disruption, signal-to-noise ratio attenuation, and sensor-tissue interface changes [99].

### Statistical analysis

All statistical analyses were performed using Python (version 3.11) with Statsmodels [83], NumPy (version 1.24) and SciPy (version 1.11) for numerical computations, custom implementations of random-effects meta-analysis using the DerSimonian-Laird (DL) estimator as DL is one of the most widely used methods for random-effects meta-analysis [84]. The I² statistic was used to quantify heterogeneity. Subgroup analyses were performed for device type, reference standard level, and study duration. Meta-regression was used to explore the association between device type and study duration, with separate regressions performed for each device type and a full interaction model to test for differential effects. The DL estimator was used for the leave-one-out sensitivity analysis for computational efficiency. Publication bias was assessed using Egger’s test and Begg’s test, with visual inspection of funnel plots [85]. All statistical tests were two-sided, with a significance threshold of p < 0.05 unless otherwise stated. Quality assessment was performed using the QUADAS-2 tool [86], and certainty of evidence was assessed using the GRADE framework [87]. Inter-rater reliability for the QUADAS-2 assessment was evaluated using the intraclass correlation coefficient (ICC, two-way random, single measures, absolute agreement). Disagreements were resolved through discussion and consensus.

## Data Availability

All data supporting the findings of this study are available within the article and its Supplementary Materials.

## Code Availability

The code used for this study is available at Github.

## Ethics Statement

This study is a systematic review and meta-analysis of previously published data and did not require ethics approval.

## Acknowledgements

Amparo Guemes Gonzalez is supported by the Royal Academy of Engineering under the Research Fellowship scheme (#RF-2324-23-284) and the Rosetrees Trust Research Fellowship. Kezhi Li and this project is supported by Internal Project Fund from UCL Institute of Health Informatics; UCL-Tohoku University Strategic Partner Funds.

## Author Contributions

K.L., H.Z., and E.D. conceived the study and established the initial research framework. E.D. designed the search strategy, led the initial data collection, and conducted preliminary analyses. H.Z. designed the updated analytical methodology and conducted the formal statistical analysis. H.Z. and E.D. carried out the systematic literature screening and eligibility assessment. K.T. and A.G. provided technical guidance on sensing physics and strategic advice on glucose monitoring performance metrics. S.E.B. provided clinical oversight and validated the medical relevance of the findings. Z.G. performed technical validation of the results and assisted in data visualisation. H.Z. wrote the original draft of the manuscript, and all other authors contributed to the critical revision of the manuscript for important intellectual content. K.L. supervised the project and serves as the guarantor.

## Supplementary Materials

### S1: Search Strategy

The following search strategies were applied to three electronic databases. The search was restricted to studies published between January 1, 2020, and November 30, 2025. Each database uses a different syntax and field structure, which required minor adaptations of the core search strategy while maintaining equivalent conceptual coverage. Review articles were excluded using database-specific publication type filters.

#### PubMed (Medline)

(glucometer[Title/Abstract] OR CGM[Title/Abstract] OR SMBG[Title/Abstract] OR “glucose monitor*”[Title/Abstract]) AND (“invasive”[Title/Abstract] OR “non-invasive”[Title/Abstract] OR transdermal[Title/Abstract] OR implantable[Title/Abstract] OR biosensors[Title/Abstract] OR CLC[Title/Abstract] OR “closed loop monitor”[Title/Abstract]) AND (diabet*[Title/Abstract] OR T1D[Title/Abstract] OR T2D[Title/Abstract]) NOT (vitro[Title/Abstract] OR Mice[Title/Abstract] OR Rat*[Title/Abstract] OR Rabbit*[Title/Abstract] OR Dog*[Title/Abstract] OR animal*[Title/Abstract] OR Injection[Title/Abstract] OR Intravenous[Title/Abstract] OR Bacteria[Title/Abstract] OR transfect*[Title/Abstract] OR plant*[Title/Abstract] OR Cells[Title/Abstract]) NOT “review”[Publication Type]

#### Web of Science (Core Collection)

TS=(glucometer OR CGM OR SMBG OR “glucose monitor*”) AND TS=(invasive OR “non-invasive” OR transdermal OR implantable OR biosensors OR CLC OR “closed loop monitor”) AND TS=(diabet* OR T1* OR T2*) NOT TS=(vitro OR Mice OR Rat* OR Rabbit* OR Dog* OR animal* OR Injection OR Intravenous OR Bacteria OR transfect* OR plant* OR Cells)

#### Scopus

TITLE-ABS-KEY(glucometer OR CGM OR SMBG OR “glucose monitor*”) AND TITLE-ABS-KEY(invasive OR “non-invasive” OR transdermal OR implantable OR biosensors OR CLC OR “closed loop monitor”) AND TITLE-ABS-KEY(diabet* OR T1* OR T2*) AND NOT TITLE-ABS-KEY(vitro OR Mice OR Rat* OR Rabbit* OR Dog* OR animal* OR Injection OR Intravenous OR Bacteria OR transfect* OR plant* OR Cells)

Note: The search strategies differ across the three databases because each database uses a distinct query syntax and indexing system. PubMed uses Title/Abstract fields, Web of Science uses Topic Search (TS=), and Scopus uses TITLE-ABS-KEY. Despite these syntactic differences, the conceptual search scope is equivalent across all three databases. The electronic database search was supplemented by a manual review of reference lists from included articles and relevant systematic reviews.

### S2: QUADAS-2 Quality Assessment

The QUADAS-2 tool [86] was applied to assess the risk of bias and applicability concerns across four domains: (D1) Patient Selection, (D2) Index Test, (D3) Reference Standard, and (D4) Flow and Timing. Each domain was rated as Low Risk, Unclear Risk, or High Risk of bias based on the signaling questions defined in the QUADAS-2 framework.

Inter-rater reliability was assessed using the intraclass correlation coefficient (ICC, two-way random, single measures, absolute agreement). The overall ICC was 0.85 (95% CI: 0.80–0.89), with a percentage agreement of 85.5% and Cohen’s kappa of 0.76, indicating excellent inter-rater consistency. Per-domain results were as follows: D1 (Patient Selection): ICC = 0.87 (95% CI: 0.77–0.93); D2 (Index Test): ICC = 0.89 (95% CI: 0.79–0.94); D3 (Reference Standard): ICC = 0.82 (95% CI: 0.68–0.90); D4 (Flow and Timing): ICC = 0.74 (95% CI: 0.57–0.86). Disagreements between the two assessors occurred in 22 of 152 individual domain ratings (14.5%), predominantly in the D3 (Reference Standard) and D4 (Flow and Timing) domains, and were resolved through discussion and consensus. Domains: D1 = Patient Selection; D2 = Index Test; D3 = Reference Standard; D4 = Flow & Timing. Ratings: Low (green) = low risk of bias; Unclear (yellow) = unclear risk of bias; High (red) = high risk of bias.

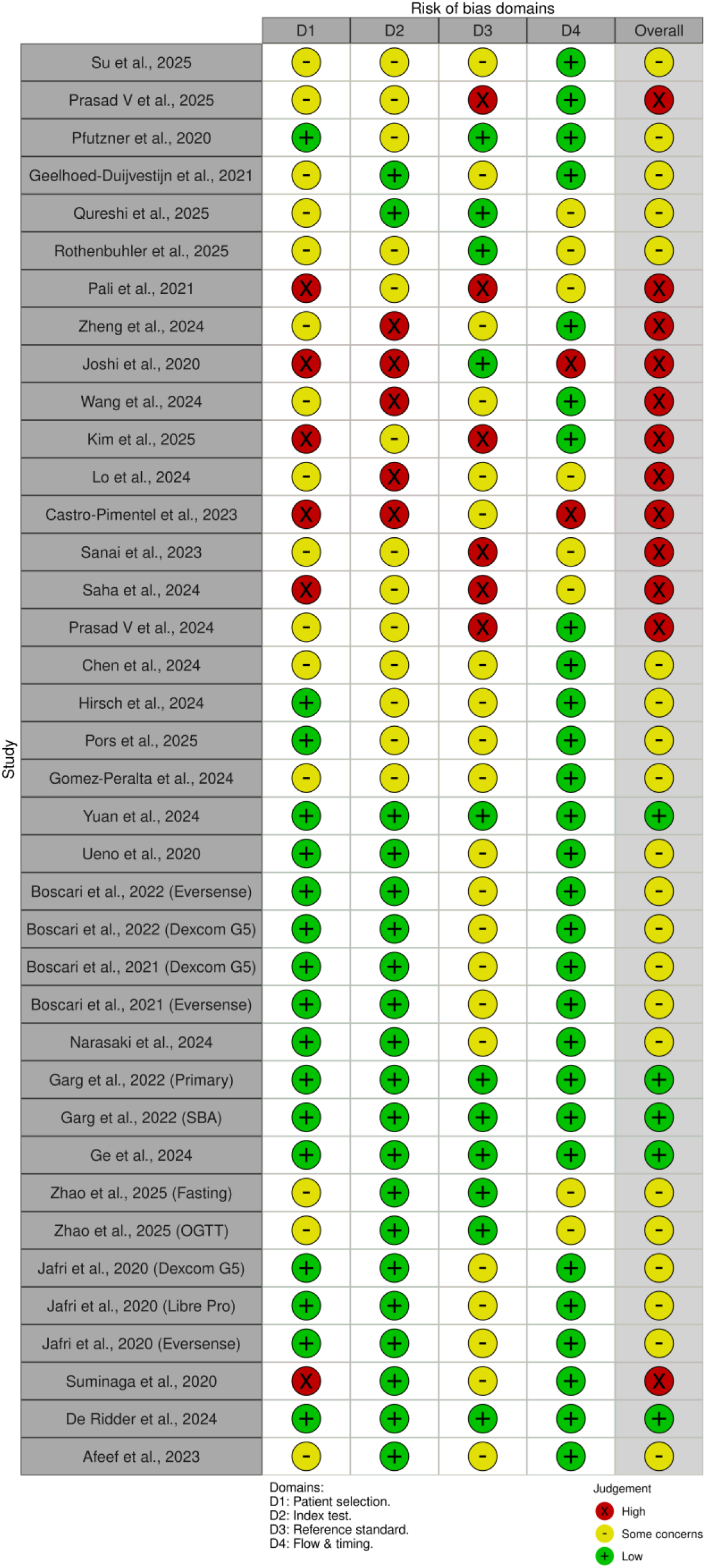

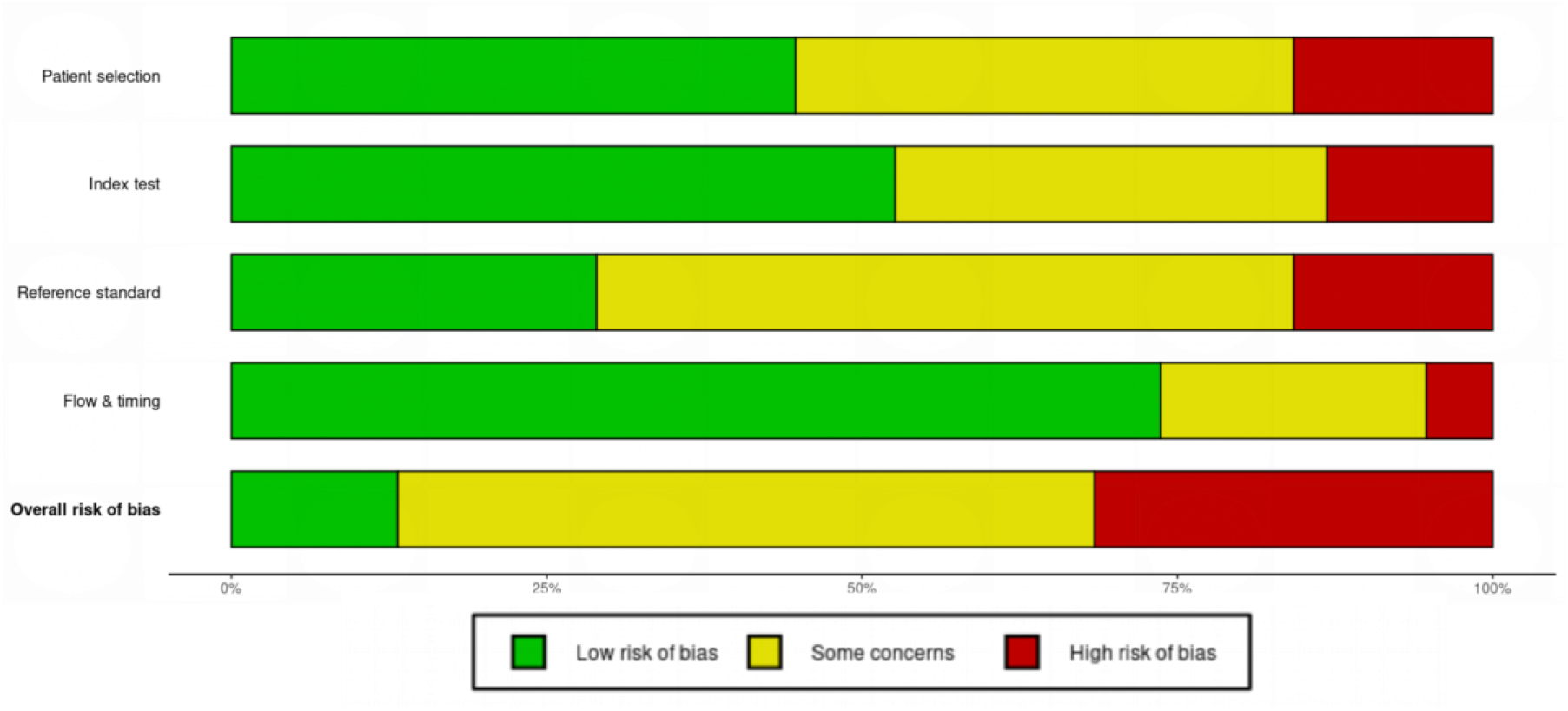

### S3: Summary of Findings and Certainty of Evidence (GRADE)

The GRADE framework [87] was used to assess the certainty of evidence for the primary outcomes. The overall certainty was rated as Very Low for all outcomes in an exploratory adapted GRADE assessment. The standard GRADE framework was developed for intervention effects and diagnostic test accuracy (sensitivity/specificity); its application to pooled MARD, a continuous measurement performance metric, represents a non-standard use. With this caveat, evidence was downgraded due to: (1) high risk of bias in the majority of included studies (downgraded one level); (2) very serious inconsistency (I² = 95.2%; downgraded two levels); (3) serious indirectness (many NIGM studies used healthy volunteers rather than the target population with diabetes; downgraded one level); (4) serious imprecision (wide confidence intervals and small cumulative sample sizes; downgraded one level); and (5) high suspicion of publication bias (Egger’s test p = 0.0002; downgraded one level).

### S4: Leave-One-Out Sensitivity Analysis

The leave-one-out sensitivity analysis was performed by sequentially omitting each of the 38 cohorts and recalculating the pooled MARD estimate using the DerSimonian-Laird random-effects model. The results demonstrate that the pooled estimate was highly consistent, with values ranging from 10.63% to 11.14% regardless of which study was omitted. No single study had a disproportionate influence on the overall result. The DL estimator was selected as it is the most widely used method in medical meta-analyses. A sensitivity analysis using the more conservative REML estimator was also performed and yielded similar results, confirming the consistency of our findings.

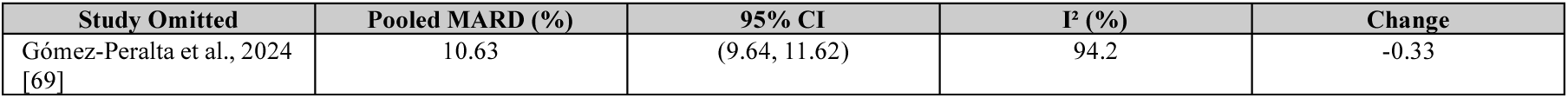
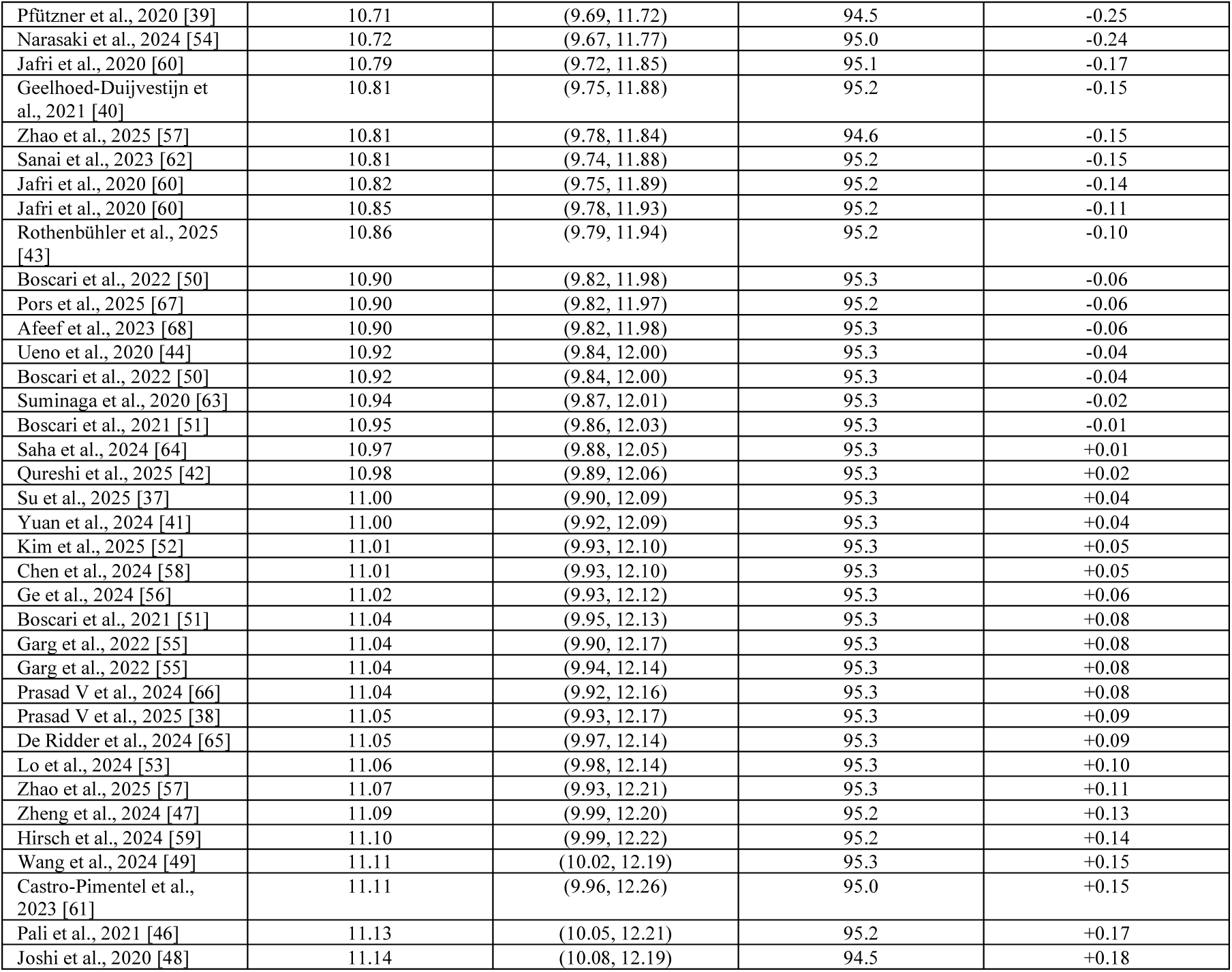

**Note: The pooled MARD estimate remained highly stable (range: 10.63–11.14%) regardless of which study was omitted, indicating consistency of the main findings. No single study had a disproportionate influence on the overall result.**

### S5: Complete Characteristics of Included Studies

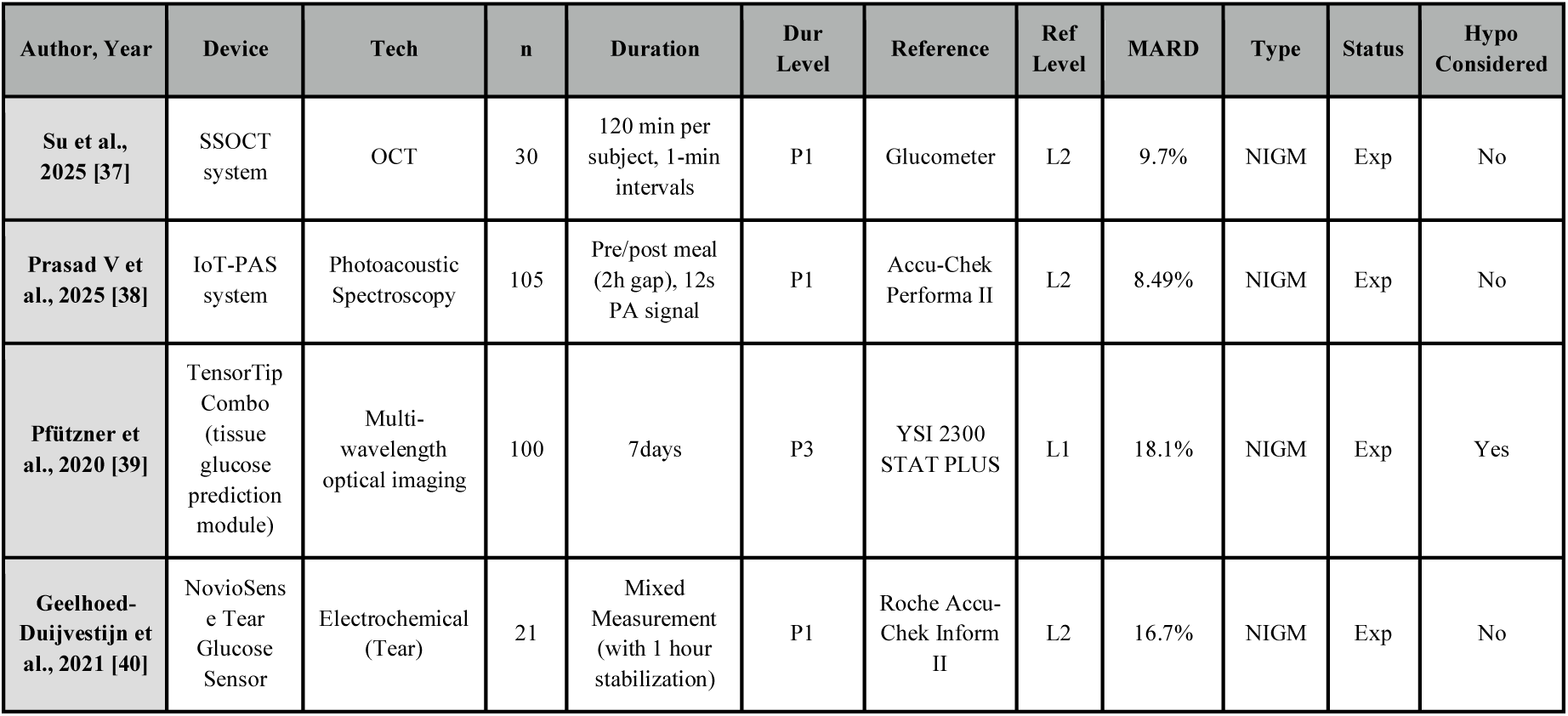
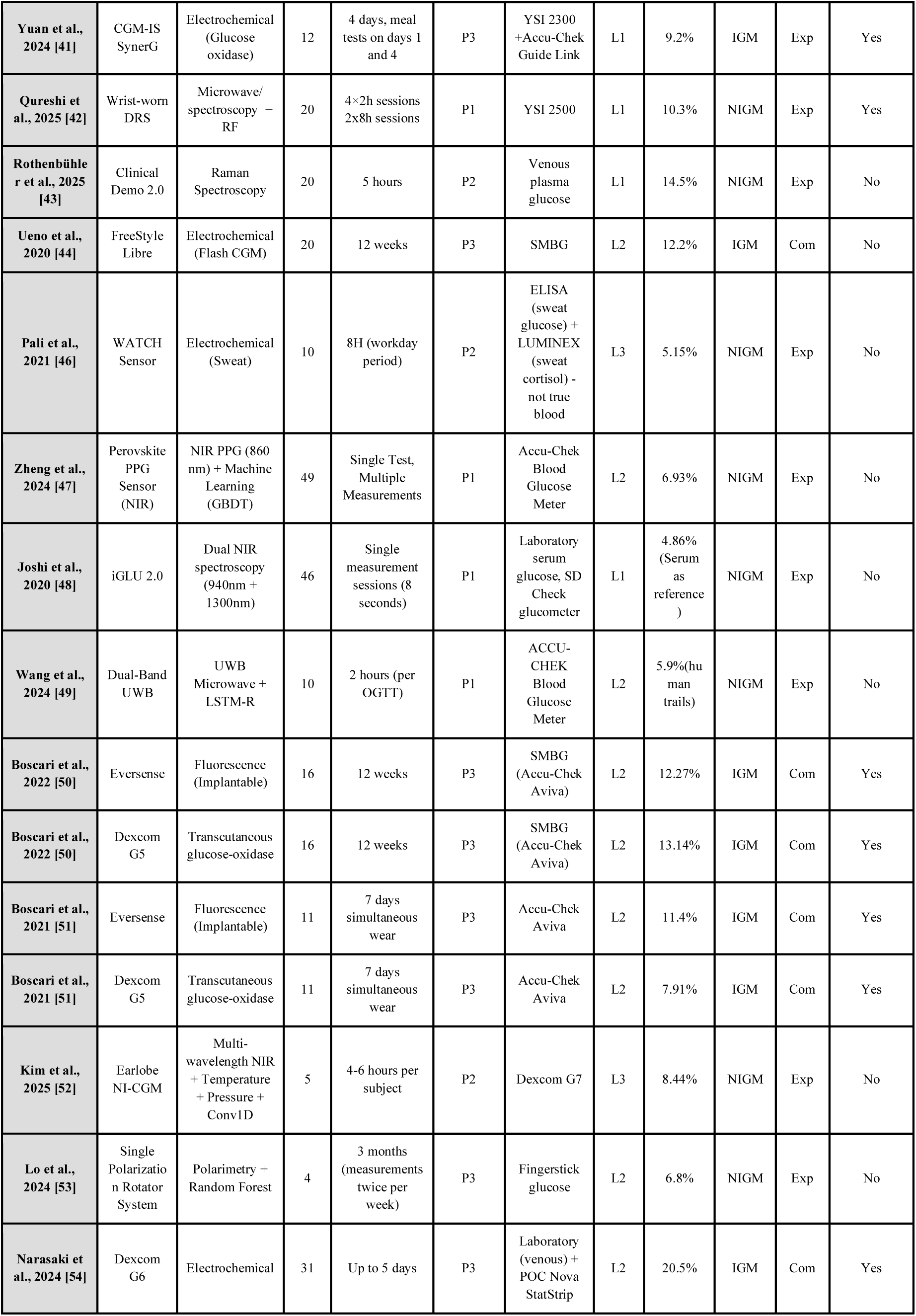
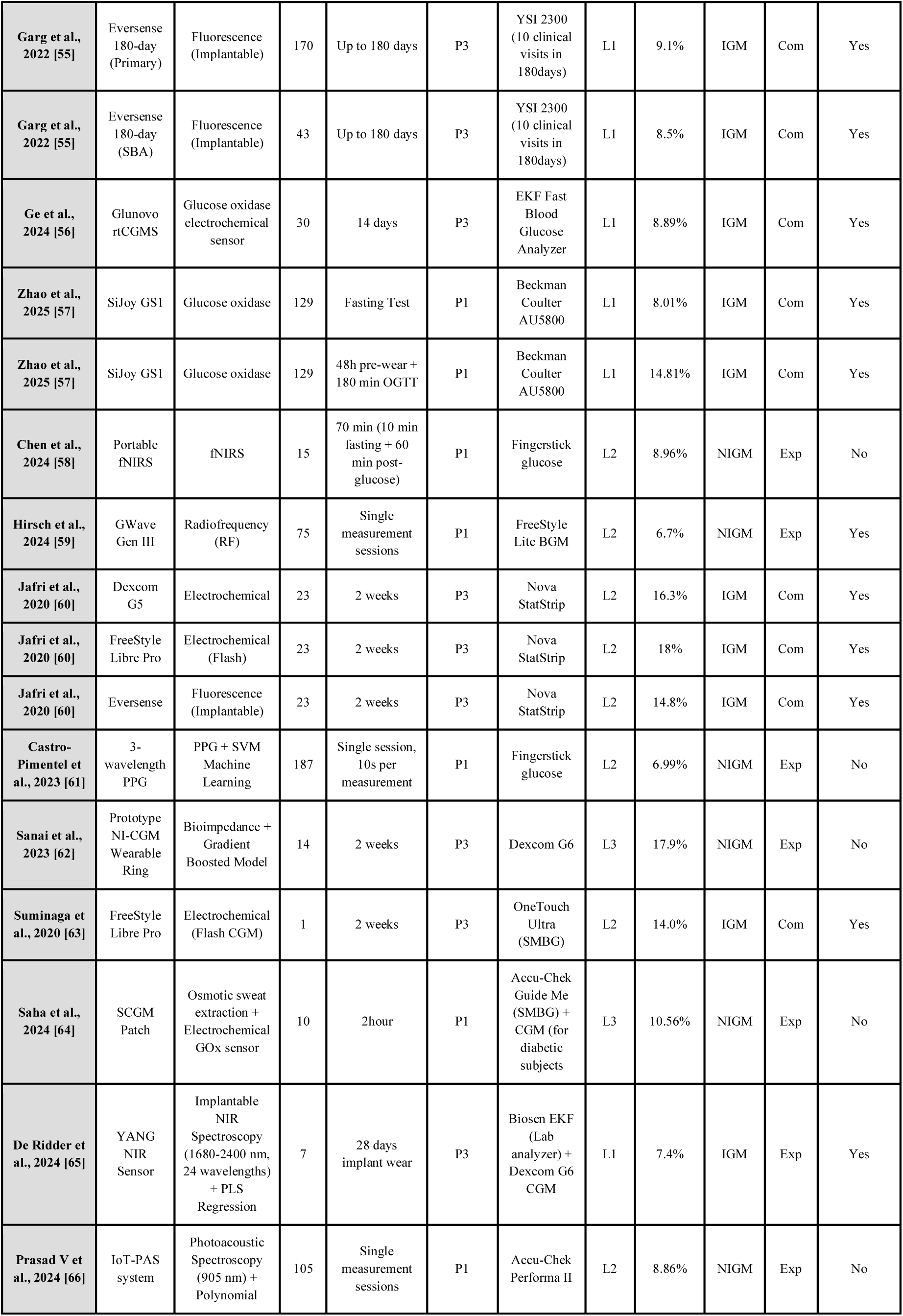
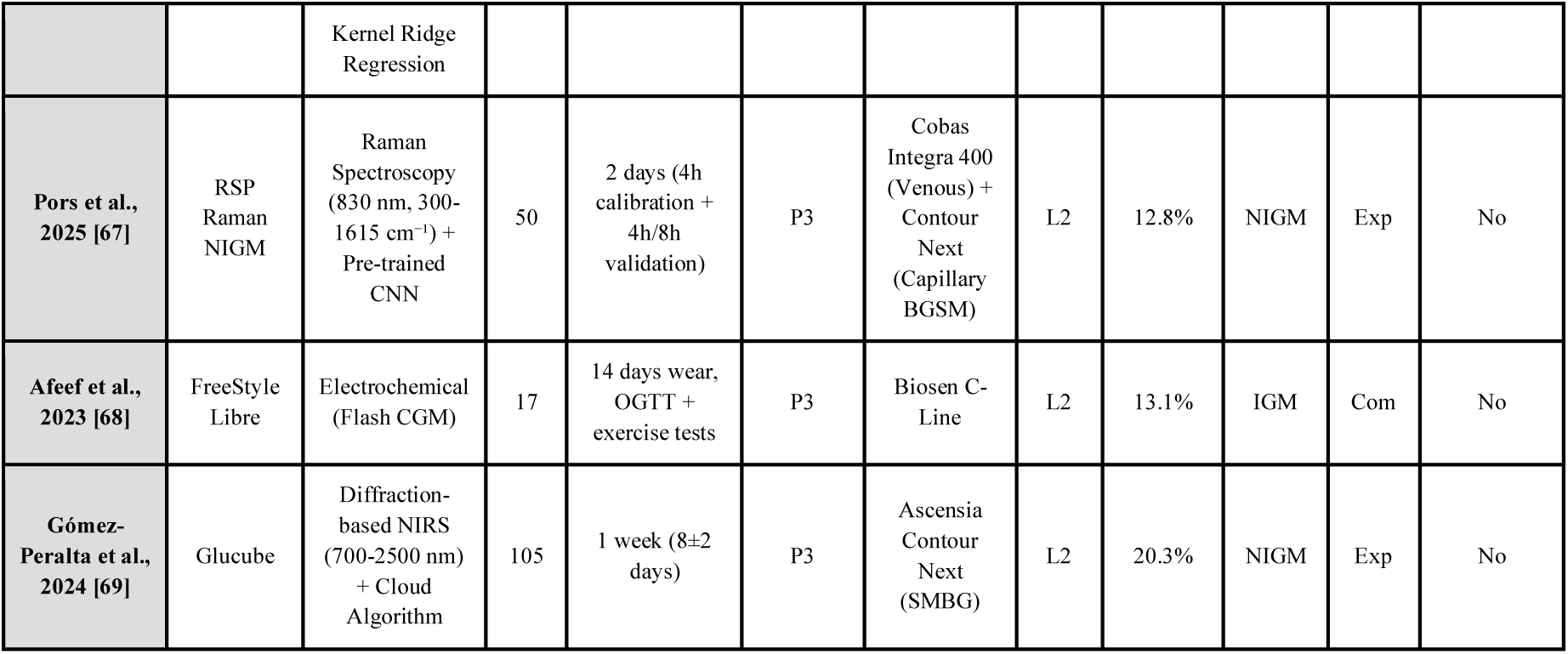

This table includes selection criteria, P1–P3 duration classification, L1–L3 reference category marks, and abbreviations for all 38 cohorts from 32 studies.

**Supplementary Figure S1.**
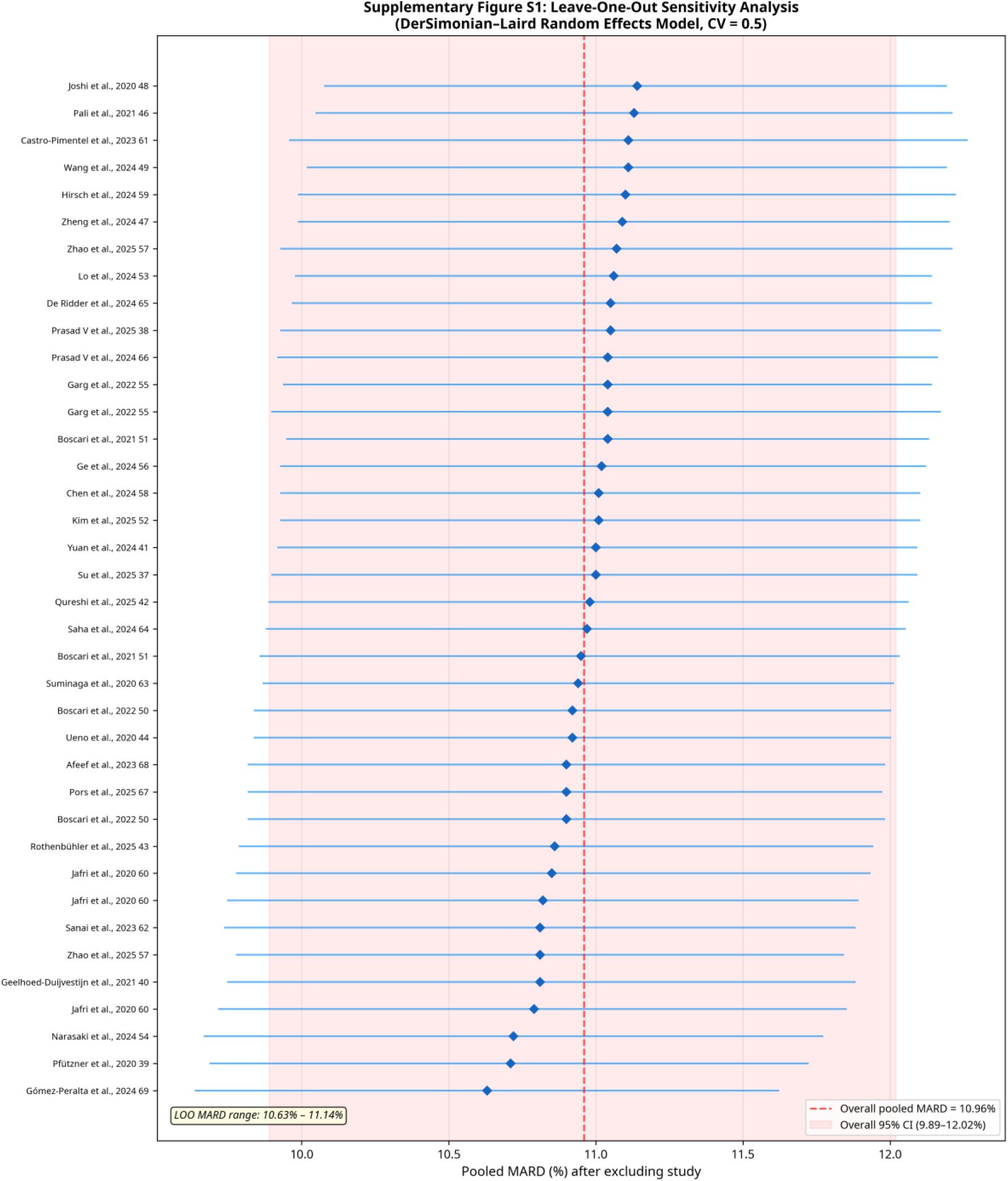
Leave-One-Out Sensitivity Analysis. Each row shows the pooled MARD estimate (red dot) and 95% confidence interval (horizontal line) after omitting the named study. The blue dashed line indicates the overall pooled MARD (10.96%), and the blue shaded band represents the overall 95% CI (9.89–12.02%). The pooled estimate remained stable (range: 10.63–11.14%) regardless of which study was omitted, indicating consistency of the main findings.

To investigate the accuracy-regulatory gap—specifically whether lower-quality reference standards produce less reliable accuracy estimates—we stratified studies by reference standard quality (Supplementary Figure S2). For NIGM devices, no significant difference in MARD was observed based on reference standard (p = 0.49). The pooled MARD was 11.9% (95% CI: 4.6–19.2%) for Level 1, 9.9% (95% CI: 8.2–11.6%) for Level 2, and 10.5% (95% CI: 5.3–15.7%) for Level 3. In contrast, for iCGM devices, the reference standard showed a non-significant association with MARD (β = 4.55, p = 0.09), with Level 1 references associated with a lower MARD (9.4%; 95% CI: 7.8–11.0%) compared to Level 2 (14.0%; 95% CI: 12.1–15.9%). This asymmetry suggests that iCGM accuracy estimates are sensitive to the precision of the reference standard, while NIGM accuracy estimates are dominated by other factors, most prominently study duration.

**Supplementary Figure S2.**
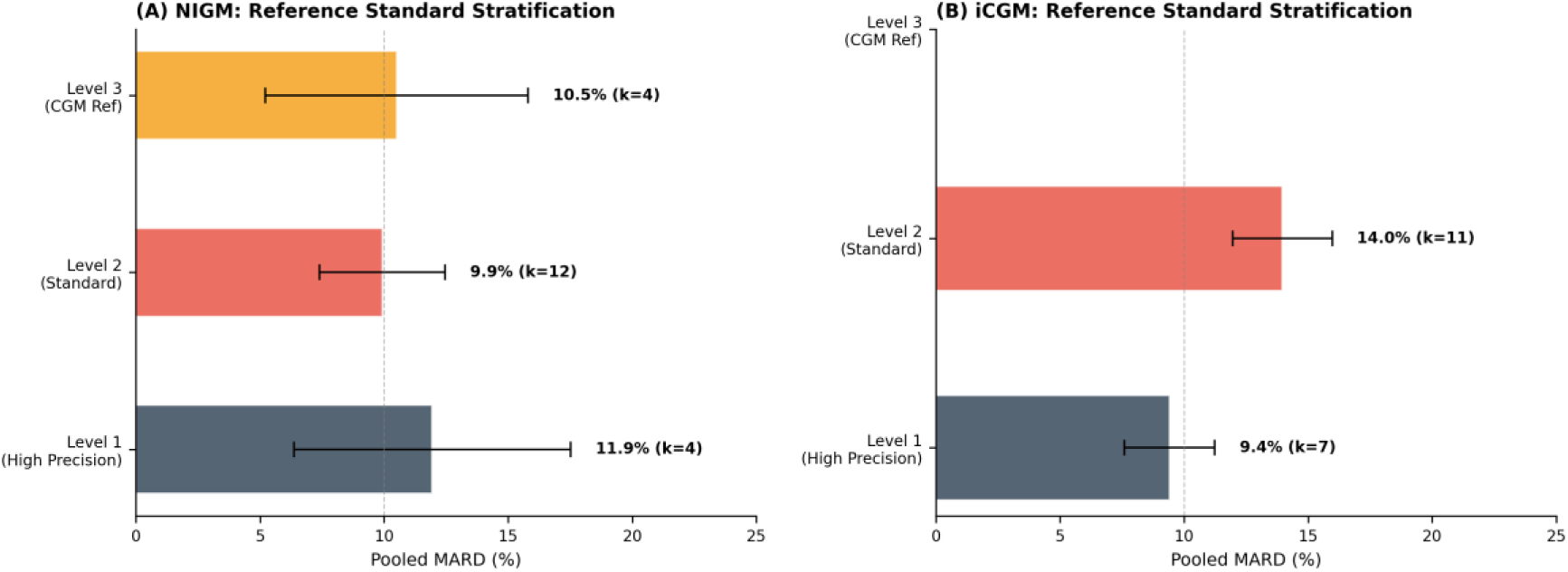
Reference to Standard Stratification. Pooled MARD for (A) NIGM and (B) iCGM cohorts, stratified by the quality of the reference standard. For iCGM, Level 1 references were numerically associated with lower MARD, though not reaching statistical significance (p = 0.09), while no significant effect was observed for NIGM (p = 0.49).

Egger’s test indicated significant asymmetry in the funnel plot (p = 0.0002), and Begg’s test was also significant (τ = 0.41, p = 0.0003), suggesting the presence of publication bias (Supplementary Figure S3). Studies with smaller sample sizes tended to report lower (more favourable) MARD values, consistent with selective publication of positive results.

**Supplementary Figure S3.**
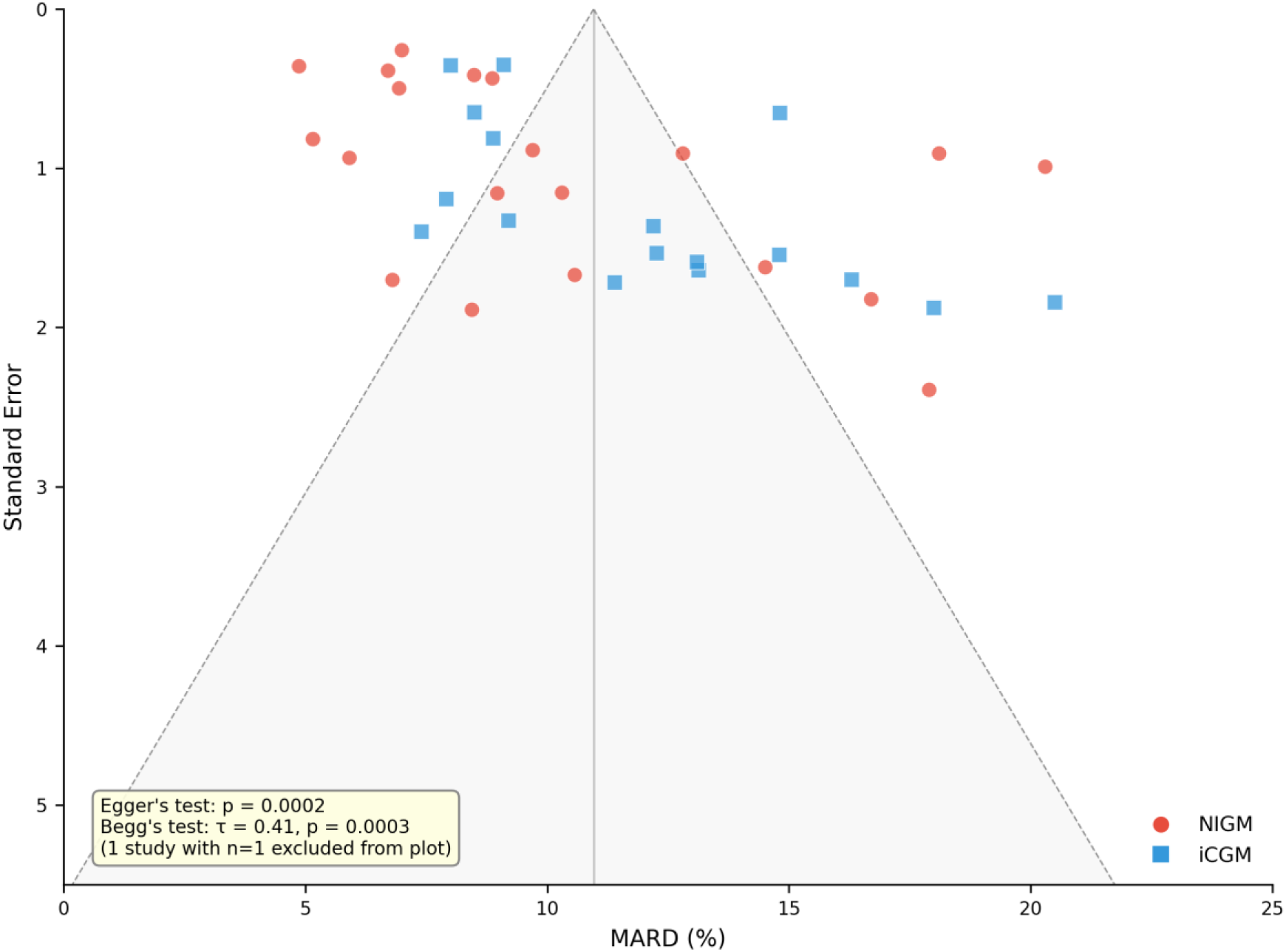
Funnel Plot for Publication Bias Assessment. The plot of MARD against standard error shows asymmetry, confirmed by Egger’s test (p = 0.0002) and Begg’s test (τ = 0.41, p = 0.0003), suggesting that smaller studies with more favourable results are more likely to be published.

### S6: PRISMA 2020 Checklist

The following checklist documents compliance with the PRISMA 2020 reporting guidelines [82]

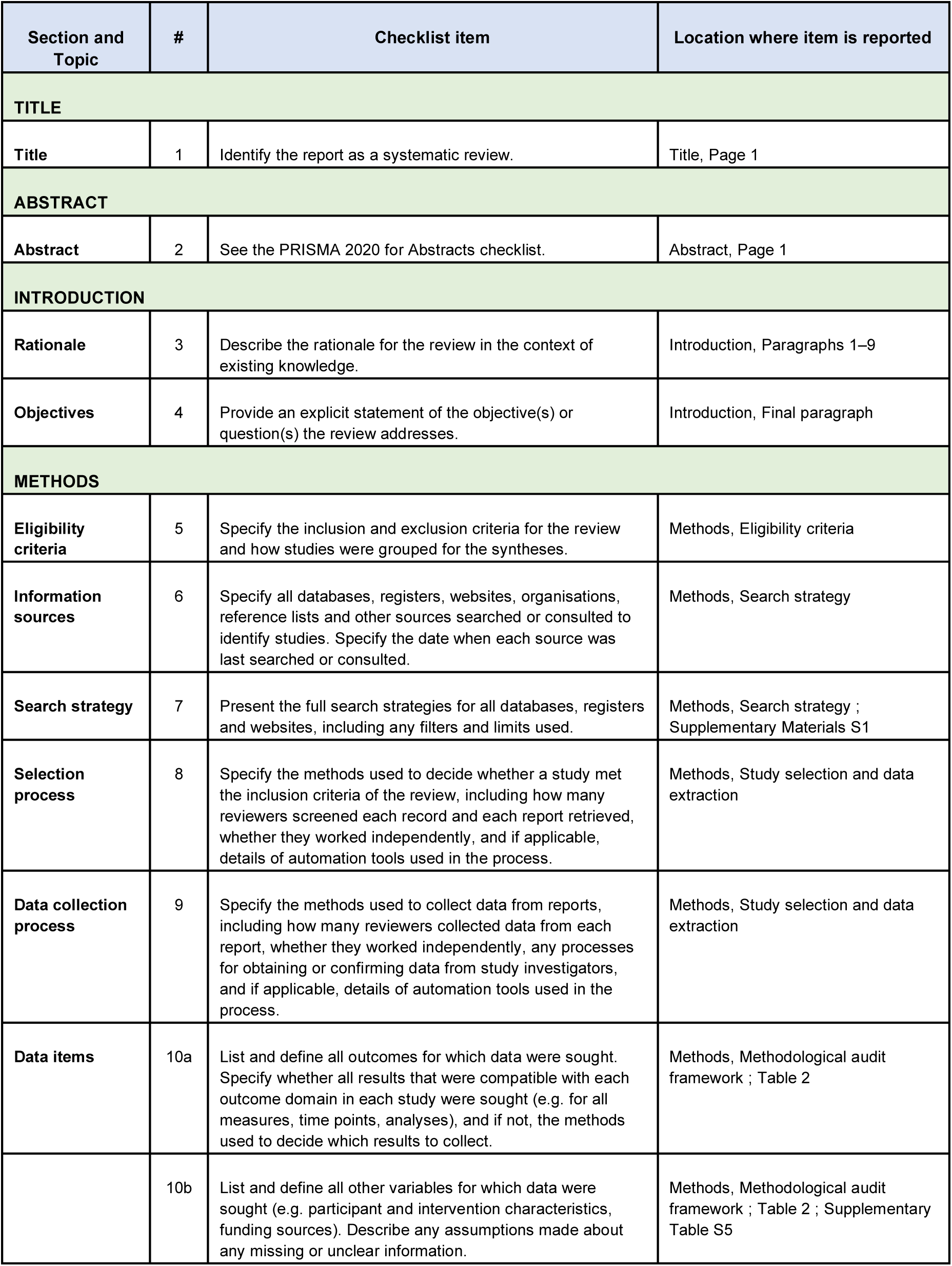
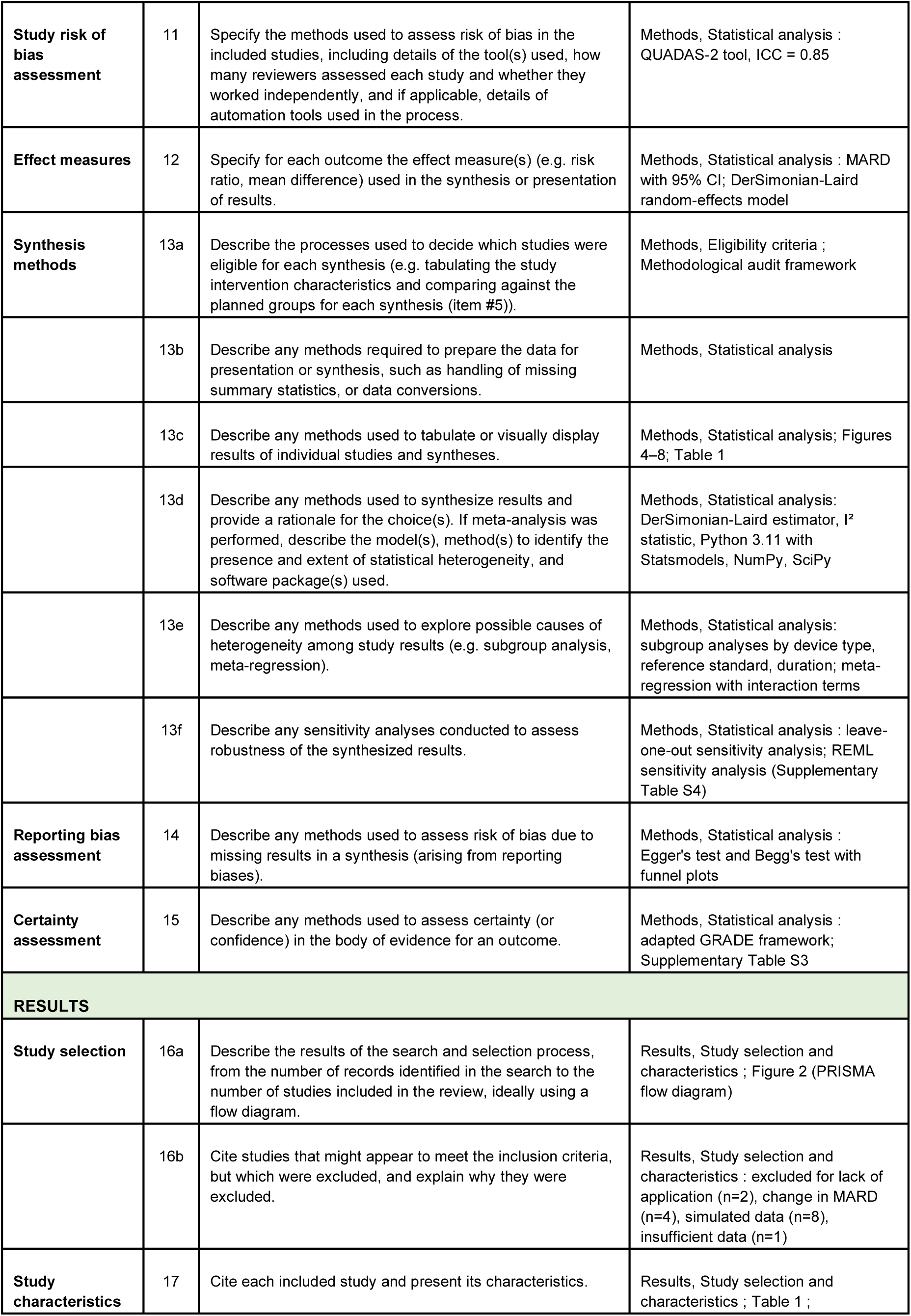
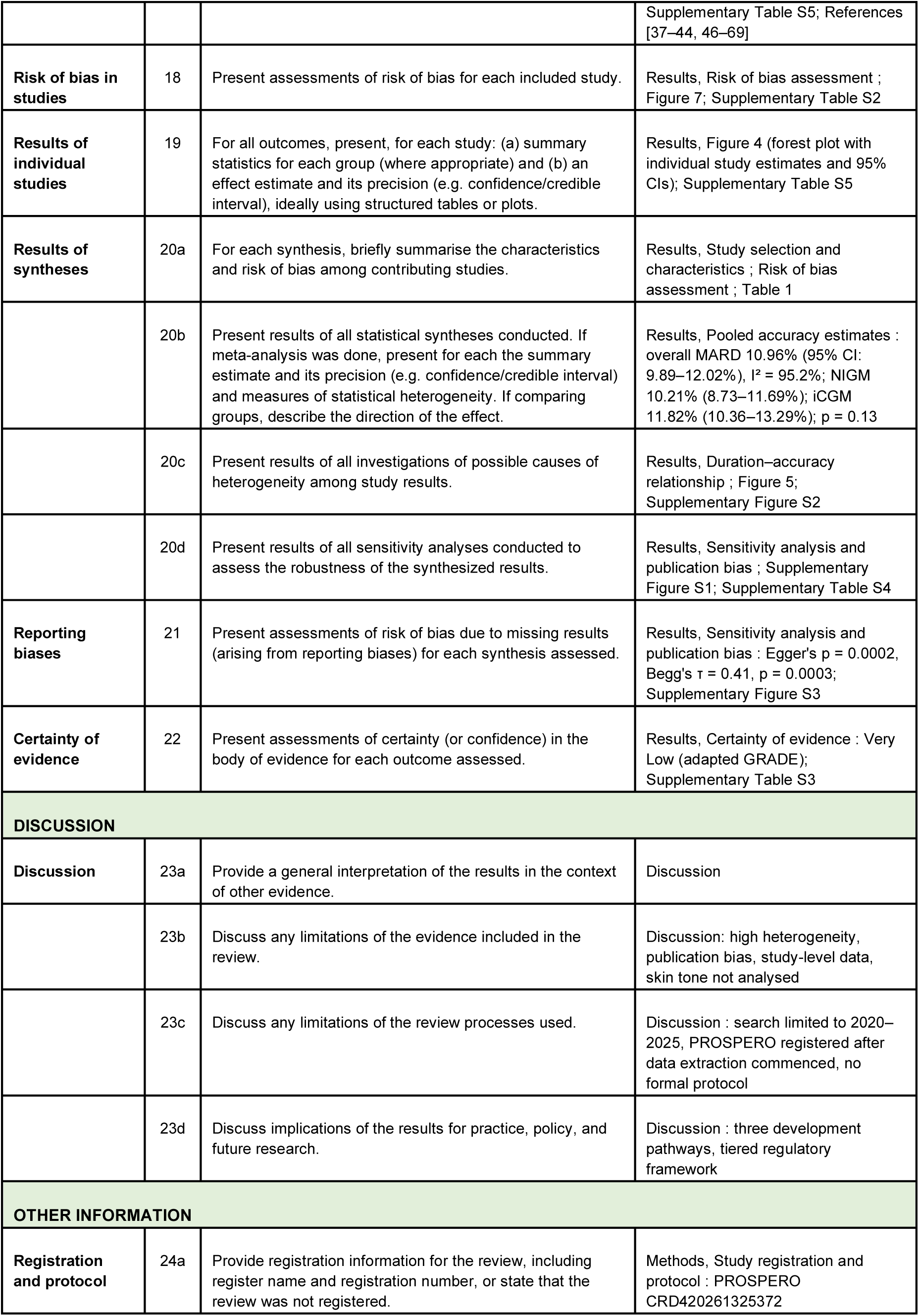
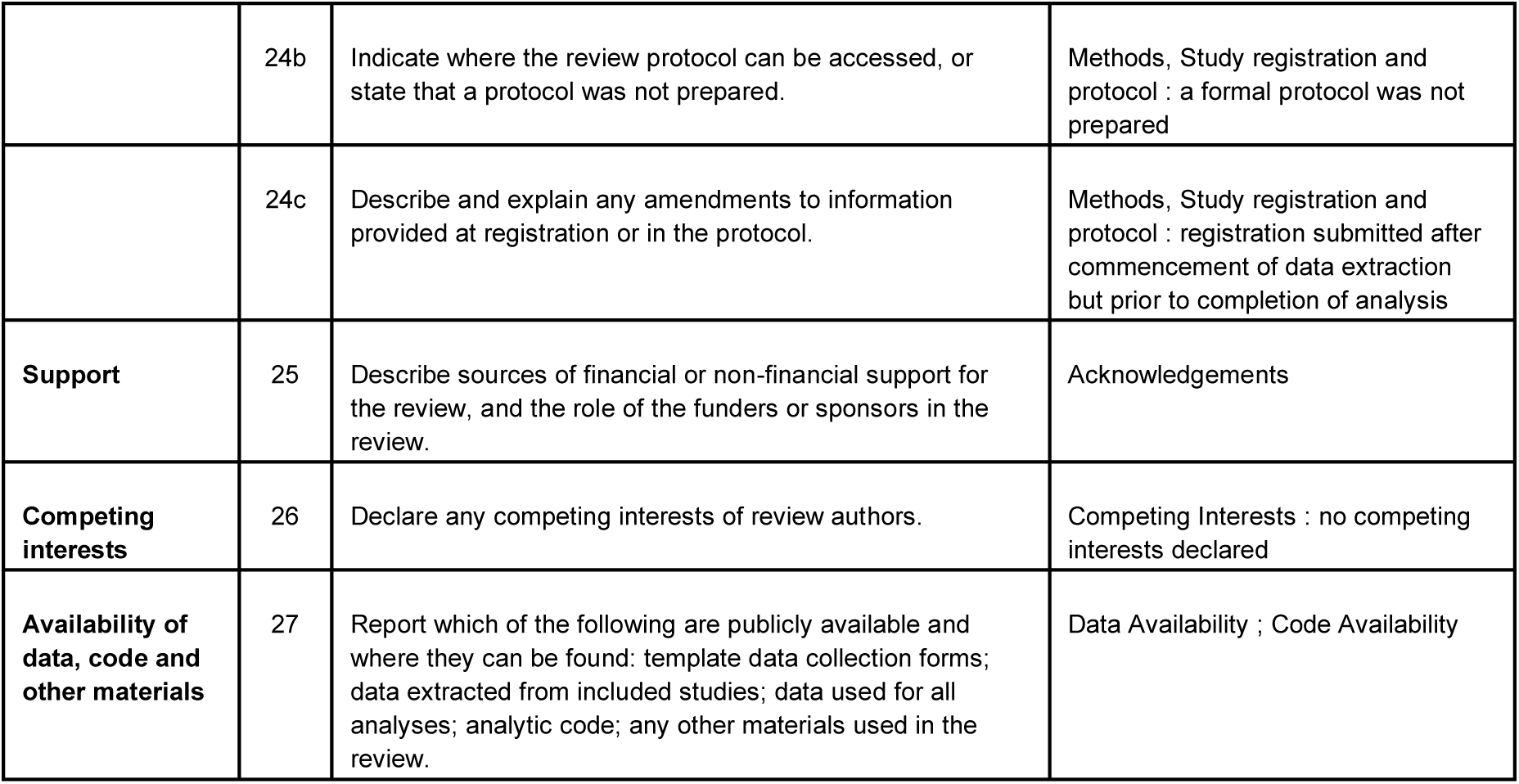

## References

[1] International Diabetes Federation. IDF Diabetes Atlas, 10th edn. Brussels, Belgium: International Diabetes Federation, 2021.

[2] American Diabetes Association. Standards of Care in Diabetes—2023. Diabetes Care 46 (Suppl. 1), S1–S291 (2023).

[3] Beck, R. W. et al. Effect of continuous glucose monitoring on glycemic control in adults with type 1 diabetes using insulin injections: the DIAMOND randomised clinical trial. JAMA 317, 371–378 (2017).

[4] Battelino, T. et al. Clinical targets for continuous glucose monitoring data interpretation: recommendations from the international consensus on time in range. Diabetes Care 42, 1593–1603 (2019).

[5] Danne, T. et al. International consensus on use of continuous glucose monitoring. Diabetes Care 40, 1631–1640 (2017).

[6] Heinemann, L. et al. Real-time continuous glucose monitoring in adults with type 1 diabetes and impaired hypoglycaemia awareness or severe hypoglycaemia treated with multiple daily insulin injections (HypoDE): a multicentre, randomised controlled trial. Lancet 391, 1367–1377 (2018).

[7] Lind, M. et al. Continuous glucose monitoring vs conventional therapy for glycemic control in adults with type 1 diabetes treated with multiple daily insulin injections: The GOLD Randomised Clinical Trial. JAMA 317, 379–387 (2017).

[8] The Diabetes Control and Complications Trial Research Group. The effect of intensive treatment of diabetes on the development and progression of long-term complications in insulin-dependent diabetes mellitus. N. Engl. J. Med. 329, 977–986 (1993).

[9] ISO 15197:2013. In vitro diagnostic test systems—Requirements for blood-glucose monitoring systems for self-testing in managing diabetes mellitus. International Organisation for Standardisation, 2013.

[10] Kovatchev, B. P. et al. Evaluating the accuracy of continuous glucose-monitoring sensors: continuous glucose-error grid analysis illustrated by TheraSense Freestyle Navigator data. Diabetes Care 27, 1922–1928 (2004).

[11] Rodbard, D. Continuous glucose monitoring: a review of successes, challenges, and opportunities. Diabetes Technol. Ther. 18 (Suppl. 2), S3–S13 (2016).

[12] Cappon, G. et al. Continuous glucose monitoring sensors for diabetes management: a review of technologies and applications. Diabetes Metab. J. 43, 383–397 (2019).

[13] Heinemann, L. & Freckmann, G. CGM versus FGM; or, continuous glucose monitoring is not flash glucose monitoring. J. Diabetes Sci. Technol. 9, 947–950 (2015).

[14] Garg, S. K. et al. Accuracy and safety of Dexcom G7 continuous glucose monitoring in adults with diabetes. Diabetes Technol. Ther. 24, 373–380 (2022).

[15] Alva, S. et al. Accuracy of the third generation of a 14-day continuous glucose monitoring system. Diabetes Ther. 14, 767–776 (2023).

[16] Christiansen, M. P. et al. A prospective multicenter evaluation of the accuracy and safety of an implanted continuous glucose sensor: the PRECISION study. Diabetes Technol. Ther. 21, 231–237 (2019).

[17] U.S. Food and Drug Administration. Special Controls for Integrated Continuous Glucose Monitoring Systems. (2018).

[18] Asarani, N. A. M. et al. Cutaneous complications with continuous or flash glucose monitoring use: systematic review of trials and observational studies. J. Diabetes Sci. Technol. 14, 328–337 (2020).

[19] Berg, A. K. et al. Skin problems associated with insulin pumps and sensors in adults with type 1 diabetes: a cross-sectional study. Diabetes Technol. Ther. 20, 475–482 (2018).

[20] Vashist, S. K. Continuous glucose monitoring systems: a review. Diagnostics 3, 385–412 (2013).

[21] Vashist, S. K. Non-invasive glucose monitoring technology in diabetes management: a review. Anal. Chim. Acta 750, 16–27 (2012).

[22] Tang, L. et al. Non-invasive blood glucose monitoring technology: a review. Sensors 20, 6925 (2020).

[23] Laha, S. et al. A concise and systematic review on non-invasive glucose monitoring for potential diabetes management. Biosensors 12, 965 (2022).

[24] Heikenfeld, J. et al. Accessing analytes in biofluids for peripheral biochemical monitoring. Nat. Biotechnol. 37, 407–419 (2019).

[25] Bandodkar, A. J. & Wang, J. Non-invasive wearable electrochemical sensors: a review. Trends Biotechnol. 32, 363–371 (2014).

[26] Hanna, J. et al. Noninvasive, wearable, and tunable electromagnetic multisensing system for continuous glucose monitoring, mimicking vasculature anatomy. Sci. Adv. 6, eaba5320 (2020).

[27] Choi, H. et al. Design and in vitro interference test of microwave noninvasive blood glucose monitoring sensor. IEEE Trans. Microw. Theory Tech. 63, 3016–3025 (2015).

[28] Caduff, A. et al. Non-invasive glucose monitoring in patients with diabetes: a novel system based on impedance spectroscopy. Biosens. Bioelectron. 22, 598–604 (2006).

[29] Huang, J., Zhang, Y. & Wu, J. Review of non-invasive continuous glucose monitoring based on impedance spectroscopy. Sens. Actuators A Phys. 311, 112103 (2020).

[30] Hina, A. & Saadeh, W. Noninvasive blood glucose monitoring systems using near-infrared technology—a review. Sensors 22, 4855 (2022).

[31] Villena Gonzales, W., Mobashsher, A. T. & Abbosh, A. The progress of glucose monitoring—a review of invasive to minimally and non-invasive techniques, devices and sensors. Sensors 19, 800 (2019).

[32] U.S. Food and Drug Administration. Do not use smartwatches or smart rings to measure blood glucose levels: FDA Safety Communication. (2024).

[33] Shokrekhodaei, M. & Quinones, S. Review of non-invasive glucose sensing techniques: optical, electrical and breath acetone. Sensors 20, 1251 (2020).

[34] Sim, J. Y. et al. In vivo microscopic photoacoustic spectroscopy for non-invasive glucose monitoring invulnerable to skin secretion products. Sci. Rep. 8, 1059 (2018).

[35] Lindner, N. et al. Non-invasive and minimally invasive glucose monitoring devices: a systematic review and meta-analysis on diagnostic accuracy of hypoglycaemia detection. Syst. Rev. 10, 145 (2021).

[36] Lin, T., Gal, A., Mayzel, Y., Horman, K. & Bahartan, K. Non-invasive glucose monitoring: a review of challenges and recent advances. Curr. Trends Biomed. Eng. Biosci. 6, 555696 (2017).

[37] Su, Y. et al. Three-dimensional correlation method for non-invasive blood glucose monitoring with optical coherence tomography. Opt. Lasers Eng. 186, 108821 (2025).

[38] Prasad V, P. N. S. B. S. V. et al. An advanced IoT-based non-invasive in vivo blood glucose estimation exploiting photoacoustic spectroscopy with SDNN architecture. Sens. Actuators A Phys. 387, 116391 (2025).

[39] Pfützner, A. et al. System accuracy assessment of a combined invasive and noninvasive glucometer. J. Diabetes Sci. Technol. 14, 575–581 (2020).

[40] Geelhoed-Duijvestijn, P. H. et al. Performance of the Prototype NovioSense Noninvasive Biosensor for Tear Glucose in Type 1 Diabetes. J. Diabetes Sci. Technol. 15, 1320–1325 (2021).

[41] Yuan, C. Y. et al. Combining an Electrochemical Continuous Glucose Sensor With an Insulin Delivery Cannula: A Feasibility Study. J. Diabetes Sci. Technol. 18, 1273–1280 (2024).

[42] Qureshi, M. R. A. et al. Using artificial intelligence to improve the accuracy of a wrist-worn, noninvasive glucose monitor: a pilot study. J. Diabetes Sci. Technol. 19, 1546–1553 (2025).

[43] Rothenbühler, M. et al. A Prospective Pilot Study Demonstrating Noninvasive Calibration-Free Glucose Measurement. J. Diabetes Sci. Technol. 10.1177/19322968251313811 (2025).

[44] Ueno, K. et al. Patient satisfaction and clinical efficacy of flash glucose monitoring in patients with type 1 diabetes: a prospective, single-center, single-arm study. Diabetes Ther. 11, 1883–1890 (2020).

[45] Pfützner, A. et al. Evaluation of the Non-Invasive Glucose Monitoring Device GlucoTrack in Patients with Type 2 Diabetes and Subjects with Prediabetes. J. Diabetes Treat. 1, 1070 (2019).

[46] Pali, M., et al. Tracking metabolic responses based on macronutrient consumption: a comprehensive study to continuously monitor and quantify dual markers (cortisol and glucose) in human sweat using WATCH sensor. Bioeng. Transl. Med. 6, e10241 (2021).

[47] Zheng, Y. et al. Highly sensitive perovskite photoplethysmography sensor for blood glucose sensing using machine learning techniques. Adv. Sci. 11, e2405681 (2024).

[48] Joshi, A. M., Jain, P., Mohanty, S. P. & Agrawal, N. iGLU 2.0: a new wearable for accurate non-invasive continuous serum glucose measurement in IoMT framework. IEEE Trans. Consum. Electron. 66, 327–335 (2020).

[49] Wang, Z. et al. Noninvasive Intelligent Blood Glucose Monitoring on Fingertip Using Dual-Band Fusion and LSTM-R Network. IEEE Sens. J. 24, 3465–3476 (2024).

[50] Boscari, F. et al. Implantable and transcutaneous continuous glucose monitoring system: a randomised cross over trial comparing accuracy, efficacy and acceptance. J. Endocrinol. Invest. 45, 115–124 (2022).

[51] Boscari, F. et al. Comparing the accuracy of transcutaneous sensor and 90-day implantable glucose sensor. Nutr. Metab. Cardiovasc. Dis. 31, 650–657 (2021).

[52] Kim, J. et al. Noninvasive Continuous Glucose Monitoring Using Multimodal Near-Infrared, Temperature, and Pressure Signals on the Earlobe. Biosensors 15, 406 (2025).

[53] Lo, Y. L. et al. Non-invasive glucose extraction by a single polarisation rotator system in patients with diabetes. Biomed. Opt. Express 15, 4909–4924 (2024).

[54] Narasaki, Y. et al. Accuracy of Continuous Glucose Monitoring in Hemodialysis Patients With Diabetes. Diabetes Care 47, 1922–1929 (2024).

[55] Garg, S. K. et al. Evaluation of Accuracy and Safety of the Next-Generation Up to 180-Day Long-Term Implantable Eversense Continuous Glucose Monitoring System: The PROMISE Study. Diabetes Technol. Ther. 24, 84–92 (2022).

[56] Ge, S. et al. Accuracy of a novel real-time continuous glucose monitoring system: a prospective self-controlled study in thirty hospitalized patients with type 2 diabetes. Front. Endocrinol. 15, 1374496 (2024).

[57] Zhao, W. et al. Clinical performance evaluation of the SiJoy GS1 continuous glucose monitor during oral glucose tolerance testing in healthy adults. Front. Endocrinol. 16, 1536292 (2025).

[58] Chen, J. et al. Exploratory insights into prefrontal cortex activity in continuous glucose monitoring: findings from a portable wearable functional near-infrared spectroscopy system. Front. Neurosci. 18, 1342744 (2024).

[59] Hirsch, I. B. et al. Noninvasive Real-Time Glucose Monitoring Is in the Near Future. Diabetes Technol. Ther. 26, 661–666 (2024).

[60] Jafri, R. et al. A Three-Way Accuracy Comparison of the Dexcom G5, Abbott Freestyle Libre Pro, and Senseonics Eversense Continuous Glucose Monitoring Devices in a Home-Use Study of Subjects with Type 1 Diabetes. Diabetes Technol. Ther. 22, 846–852 (2020).

[61] Castro-Pimentel, L. A., Téllez-Anguiano, A. d. C., Coronado-Reyes, O. I. & Diaz-Huerta, J. L. Three-wavelength PPG and support vector machine for non-invasive estimation of blood glucose. Opt. Quantum Electron. 55, 708 (2023).

[62] Sanai, F. et al. Evaluation of a Continuous Blood Glucose Monitor: A Novel and Non-Invasive Wearable Using Bioimpedance Technology. J. Diabetes Sci. Technol. 17, 336–344 (2023).

[63] Suminaga, K. et al. Factory-calibrated continuous glucose monitoring and capillary blood glucose monitoring in a case with insulinoma: usefulness and possible pitfall under chronic hyperinsulinemic hypoglycemia. Endocr. J. 67, 361–366 (2020).

[64] Saha, T. et al. A Passive Perspiration Inspired Wearable Platform for Continuous Glucose Monitoring. Adv. Sci. 11, e2405518 (2024).

[65] De Ridder, F. et al. Early feasibility study with an implantable near-infrared spectroscopy sensor for glucose, ketones, lactate and ethanol. PLOS ONE 19, e0301041 (2024).

[66] Prasad V, P. N. S. B. S. V. et al. Augmenting authenticity for non-invasive in vivo detection of random blood glucose with photoacoustic spectroscopy using Kernel-based ridge regression. Sci. Rep. 14, 8352 (2024).

[67] Pors, A. et al. Calibration and performance of a Raman-based device for non-invasive glucose monitoring in type 2 diabetes. Sci. Rep. 15, 10226 (2025).

[68] Afeef, S. et al. Performance of the FreeStyle Libre Flash Glucose Monitoring System during an Oral Glucose Tolerance Test and Exercise in Healthy Adolescents. Sensors 23, 4249 (2023).

[69] Gómez-Peralta, F. et al. Performance of a Non-Invasive System for Monitoring Blood Glucose Levels Based on Near-Infrared Spectroscopy Technology (Glucube®). Sensors 24, 7811 (2024).

[70] Heikenfeld, J. et al. Wearable sensors: modalities, challenges, and prospects. Lab Chip 18, 217–248 (2018).

[71] Enejder, A. M. K. et al. Raman spectroscopy for noninvasive glucose measurements. J. Biomed. Opt. 10, 031114 (2005).

[72] Maruo, K. et al. In vivo noninvasive measurement of blood glucose by near-infrared diffuse-reflectance spectroscopy. Appl. Spectrosc. 57, 1236–1244 (2003).

[73] Jacques, S. L. Optical properties of biological tissues: a review. Phys. Med. Biol. 58, R37–R61 (2013).

[74] Omer, A. E. et al. Low-cost portable microwave sensor for non-invasive monitoring of blood glucose level: novel design utilizing a four-cell CSRR hexagonal configuration. Sci. Rep. 10, 15200 (2020).

[75] Arnold, M. A. & Small, G. W. Noninvasive glucose sensing. Anal. Chem. 77, 5429–5439 (2005).

[76] Cryer, P. E. Hypoglycemia, functional brain failure, and brain death. J. Clin. Invest. 117, 868–870 (2007).

[77] Frier, B. M. Hypoglycaemia in diabetes mellitus: epidemiology and clinical implications. Nat. Rev. Endocrinol. 10, 711–722 (2014).

[78] Seaquist, E. R. et al. Hypoglycemia and diabetes: a report of a workgroup of the American Diabetes Association and the Endocrine Society. Diabetes Care 36, 1384–1395 (2013).

[79] Kaysir, M. R. et al. Photoacoustic Resonators for Non-Invasive Blood Glucose Detection Through Photoacoustic Spectroscopy: A Systematic Review. Sensors 24, 6963 (2024).

[80] Shao, J. et al. In vivo blood glucose quantification using Raman spectroscopy. PLOS ONE 7, e48127 (2012).

[81] Teymourian, H. et al. Microneedle-based detection of ketone bodies along with glucose and lactate: toward real-time continuous interstitial fluid monitoring of diabetic ketosis and ketoacidosis. Anal. Chem. 92, 2291–2300 (2020).

[82] Page, M. J., et al. The PRISMA 2020 statement: an updated guideline for reporting systematic reviews. BMJ 372, n71 (2021).

[83] Seabold, S. & Perktold, J. Statsmodels: econometric and statistical modeling with Python. in Proceedings of the 9th Python in Science Conference (SciPy 2010), 92–96 (2010).

[84] Veroniki, A. A. et al. Methods to estimate the between-study variance and its uncertainty in meta-analysis. Res. Synth. Methods 7, 55–79 (2016).

[85] Egger, M., Davey Smith, G., Schneider, M. & Minder, C. Bias in meta-analysis detected by a simple, graphical test. BMJ 315, 629–634 (1997).

[86] Whiting, P. F. et al. QUADAS-2: a revised tool for the quality assessment of diagnostic accuracy studies. Ann. Intern. Med. 155, 529–536 (2011).

[87] Guyatt, G. H. et al. GRADE: an emerging consensus on rating quality of evidence and strength of recommendations. BMJ 336, 924–926 (2008).

[88] Fraser, R. A., Walker, R. J., Campbell, J. A. et al. Integration of artificial intelligence and wearable technology in the management of diabetes and prediabetes. npj Digit. Med. 8, 687 (2025).

[89] Bent, B., Cho, P. J., Henriquez, M. et al. Engineering digital biomarkers of interstitial glucose from noninvasive smartwatches. npj Digit. Med. 4, 89 (2021).

[90] Zhu, T., Uduku, C., Li, K. et al. Enhancing self-management in type 1 diabetes with wearables and deep learning. npj Digit. Med. 5, 78 (2022).

[91] Delbeck, S., Vahlsing, T., Leonhardt, S. et al. Non-invasive monitoring of blood glucose using optical methods for skin spectroscopy—opportunities and recent advances. Anal. Bioanal. Chem. 411, 63–77 (2019).

[92] Haupt, A., Berg, B., Paschen, P. et al. The effects of skin temperature and testing site on blood glucose measurements taken by a modern blood glucose monitoring device. Diabetes Technol. Ther. 7, 597–601 (2005).

[93] Mittal, R., Koutras, N., Maya, J., Lemos, J. R. N. & Hirani, K. Blood glucose monitoring devices for type 1 diabetes: a journey from the food and drug administration approval to market availability. Front. Endocrinol. 15, 1352302 (2024).

[94] Borenstein, M. How to understand and report heterogeneity in a meta-analysis: the difference between I-squared and prediction intervals. Integr. Med. Res. 12, 101014 (2023).

[95] Choi, G. J. & Kang, H. Heterogeneity in meta-analyses: an unavoidable challenge worth exploring. Korean J. Anesthesiol. 78, 301–314 (2025).

[96] Alba, A. C. et al. High statistical heterogeneity is more frequent in meta-analysis of continuous than binary outcomes. J. Clin. Epidemiol. 70, 129–135 (2016).

[97] Cengiz, E. & Tamborlane, W. V. A tale of two compartments: interstitial versus blood glucose monitoring. Diabetes Technol. Ther. 11 (Suppl. 1), S-11–S-16 (2009).

[98] Vaddiraju, S., Burgess, D. J., Tomazos, I., Jain, F. C. & Papadimitrakopoulos, F. Technologies for Continuous Glucose Monitoring: Current Problems and Future Promises. J. Diabetes Sci. Technol. 4, 1540–1562 (2010).

[99] Caduff, A., Hirt, E., Feldman, Y., Ali, Z. & Heinemann, L. First human experiments with a novel non-invasive, non-optical continuous glucose monitoring system. Biosens. Bioelectron. 19, 209–217 (2003).

[100] Liu, H., Liu, W., Sun, C., Huang, W. & Cui, X. A review of non-invasive blood glucose monitoring through breath acetone and body surface. Sens. Actuators A: Phys. 374, 115500 (2024).

[101] Lombardi, S., Bocchi, L. & Francia, P. Photoplethysmography and artificial intelligence for blood glucose level estimation in diabetic patients: a scoping review. IEEE Access 12, 178982–178996 (2024).

[102] U.S. Food and Drug Administration. General Wellness: Policy for Low Risk Devices – Guidance for Industry and Food and Drug Administration Staff. FDA-2014-N-1039 (2026).

